# Mathematical modelling of pregnant women coinfected with HIV and ZIKV: A case study in endemic Latin American and Caribbean countries

**DOI:** 10.64898/2026.01.15.26344191

**Authors:** Sika-Rose Coffi, Jhoana P. Romero-Leiton, Idriss Sekkak, Rado Ramasy, Bouchra Nasri

## Abstract

**Background:** The dynamics of HIV and ZIKV coinfection among pregnant women remain understudied, and its impacts on neonatal health still need to be defined. This gap is particularly concerning given the significant public health risks it can cause, especially in Latin America and the Caribbean, where the Zika virus is still circulating.

**Methods:** We conducted a transversal ecological study using aggregated data from 2015 to 2023. To do so, we developed a compartmental model that included a Susceptible-Infected-Recovered (SIR) compartment for pregnant women related to HIV and ZIKV infection status, as well as SI compartments for their newborns and ZIKV-carrying mosquitoes to perform simulations representing different epidemiological scenarios. We calculated the HIV/ZIKV basic reproduction number R0. Sensitivity analysis was performed to identify the model parameter with the greatest impact on this metric. Finally, we applied personal and sexual protection, medical treatment for Zika, and antiretroviral therapy as control measures against viral infections to evaluate the most effective strategy to improve neonatal health outcomes.

**Results:** The basic reproduction number ℛ_0_ associated with HIV/ZIKV coinfection among pregnant women varied in the range [0.09, 1.29] in the countries studied. The sensitivity analysis revealed that the ℛ_0_ was most sensitive to the mosquito biting rate and the pregnant women’s death rate. ZIKV infection rate among pregnant women had a greater impact on the number of newborns with related health problems compared to HIV infection rate. The introduction of ZIKV among pregnant women was enough to cause a surge in the number of newborns with related health issues, demonstrating a greater impact than HIV on neonatal health. When control strategies were applied to the model, simulations demonstrated that their application needed to be maintained concurrently over time and that medical treatment of Zika was the measure having the least influence on the coinfection.

**Conclusion:** Since pregnant women infected with HIV are particularly vulnerable to other infections such as ZIKV, it is crucial to improve antenatal care among them. Continued monitoring and increased prevention of HIV sexual transmission is also necessary to ensure maternal and child health. In order to optimize public health interventions, it is important to extend the research to strengthen our understanding of the implications of HIV-ZIKV coinfection among pregnant women and its effects on their children.

## 1 Introduction

The Zika virus pandemic in Latin America and the Caribbean in 2015-2016 led the World Health Organization (WHO) to declare a public health emergency of international concern in February 2016 [1]. The virus demonstrated its rapid spread primarily through vectors, the *Aedes* mosquitoes, capable of transmitting Dengue and Chikungunya as well, but also by sexual transmission [2, 3]. Its monitoring and control are complicated because 80 % of infected individuals do not exhibit symptoms [2]. That research led to the identification of the congenital Zika syndrome by the Centers for Disease Control and Prevention, which includes severe developmental anomalies attributed to direct viral effects on nervous tissue in babies exposed to ZIKV during gestation [4, 5]. Amid these concerns, projections show that more than 1.3 billion new people could become at risk of infection by 2050 [6], raising concerns about a potential future Zika outbreak. Moreover, studies underlined that global warming and environmental changes may lead the United States and southern Canada to become conducive environments for *Aedes spp*. mosquitoes by 2100 [7].

Extensive research is available about ZIKV alone, but there is only limited research about ZIKV coinfection with other viruses, such as HIV, especially among pregnant women [8, 9]. From 2000 to 2020, data from the Brazilian Ministry of Health indicated that 134,328 pregnant women tested positive for HIV [10]. The women constitute a particularly vulnerable population due to their immunocompromised status, making them susceptible to secondary infections such as ZIKV. They thus require acute prenatal care, including antiretroviral treatment (ART), to reduce the risk of vertical virus transmission to their newborns [2, 11, 12]. Despite the critical implications, only a few models have been developed to address this coinfection, and none of them have focused specifically on the pregnant women population [9, 13, 14].

Furthermore, in Rio de Janeiro, from January 2015 to August 2016, 219 pregnant women from a mother-to-child transmission prevention center for HIV-positive women participated in the largest case study on ZIKV and HIV coinfection in Brazil [15]. The prevalence of neurological disorders among children exposed to ZIKV in utero was found to be 5%, but this study revealed a 12% prevalence when the newborns were exposed to both ZIKV and HIV [15]. However, only limited data is available to understand the coinfection impact on pregnant women’s immune systems and newborn’s health [16]. Given the rarity of the occurrence of the HIV-ZIKV coinfection among pregnant women and the challenges in identifying and studying such a complex event, it is difficult to constitute a cohort to highlight the dynamics of the coinfection. In fact, an international prospective cohort study in 2017 aimed to address this gap but failed to enroll pregnant women infected with ZIKV [17].

To understand and quantify the impact of the coinfection on a newborn’s health, we propose a mathematical compartmental model specifically designed to analyze the coinfection dynamics of ZIKV and HIV in pregnant women, based on the formulation in [13]. This model integrates variables such as the number of pregnant women with either or both viruses and the biological vectors’ population. The objective is to deepen our understanding of the concurrence of this coinfection and its broader implications for maternal and neonatal health. Additionally, projections realized with the model can be used as a tool for the development of effective public health strategies to mitigate the burden of this coinfection in ZIKV-endemic regions of Latin America and the Caribbean.

Our primary objective is to quantify the number of women who give birth to babies with health complications related to maternal ZIKV and HIV coinfection. Thus, we will simulate different epidemiological scenarios by varying initial conditions for HIV and ZIKV among pregnant women. Finally, we will introduce control strategies to the model to compare their effectiveness in reducing the number of altered births.

## 2 Materials and methods

### 2.1 Study design and population

We conducted an ecological cross-sectional study using aggregated national data from 2015 to 2022, based on the compartmental mathematical model designed to simulate the dynamics of HIV and ZIKV coinfections among pregnant women. The selection process of the countries included in our study was based on the prevalence of both viruses, as well as the historical significance of these viruses as reported by national health agencies and reports [18, 19]. Thus, we selected Brazil, Colombia, Bolivia, El Salvador, Guatemala, and Puerto Rico. We also used data available from a study conducted in Brazil in 2015, which provided detailed information on HIV and ZIKV coinfection rates during a period of high ZIKV transmission in the country [15], and projected these onto the 2022 population for our simulations.

### 2.2 Mathematical model formulation and assumptions

In this model, the interaction between the population of pregnant women (host) and mosquitoes (vectors) is investigated. The formulation is based on several key assumptions:

- The transmission of ZIKV among women is modelled using a susceptible-infected-recovered (SIR) structure for pregnant women; we also considered two additional compartments for those women who give birth to babies with health complications associated with HIV or zika (*P*) and without health complications (*H*) following the approach in [20]. For the mosquito compartment, a susceptible-infected (SI) structure is used. The model employs a susceptible-infected AIDS (SIA) structure regarding HIV transmission, where *A* represents the compartment for pregnant women with AIDS.
- Initially, the model does not include any intervention strategies to limit propagation of the viruses. In a later section, we discuss four potential strategies: using personal protection against mosquito bites, antiretroviral therapy (ART), personal protection against HIV, and medical treatment for zika.
- It is assumed that a completely susceptible pregnant woman can contract zika through a bite from a ZIKV-carrier mosquito with probability *β*_*hv*_, or they can contract HIV through the exchange of infected syringes, blood transfusion, or sexual contact with an individual infected with HIV at a constant rate *ϕ*.
- Since the model exclusively focuses on the population of pregnant women and women who give birth, HIV transmission is not considered through sexual contact between individuals by mass action law, but rather at a constant rate *ϕ*.
- We only consider health issues associated with HIV, ZIKV, or coinfections, such as microcephaly or Guillain-Barré syndrome; it means that susceptible and zika-recovered pregnant women give birth only to healthy babies at a rate *τ*_1_. Therefore, our model does not consider other types of health associated with those disease complications. This is because it has been proven that the number of congenital malformations and particularly microcephaly prevalence among newborns more than doubled in response to the ZIKV epidemic in the region in 2015, from 0.6 for the 2010-2014 timeframe to 8 per 10000 in 2016, demonstrating its huge impact on that rare phenomenon among the uninfected population [21].
- We assume that the rate at which pregnant women infected with ZIKV, HIV, or both give birth to healthy babies is defined by the formula *τ*_2_(1 − *ψ*) where *τ*_2_ is the rate at which infected women give birth and *ψ* is the probability of fetal malformation due to infection.

The pregnant woman population is divided into six compartments, while the mosquito population is divided into two depending on their infection status. A compartment regarding the health outcomes of newborns is also included. A nonlinear system of ordinary differential equations (ODEs) governs the transitions between compartments. The compartmental structure of the model is illustrated in Figure 1, with the dynamic variables, initial conditions, and fixed parameters listed in Tables 1. The model categorizes the population of women at any given time *t*, denoted by *N* (*t*), into different compartments that can be found in 1.

**Table 1:**
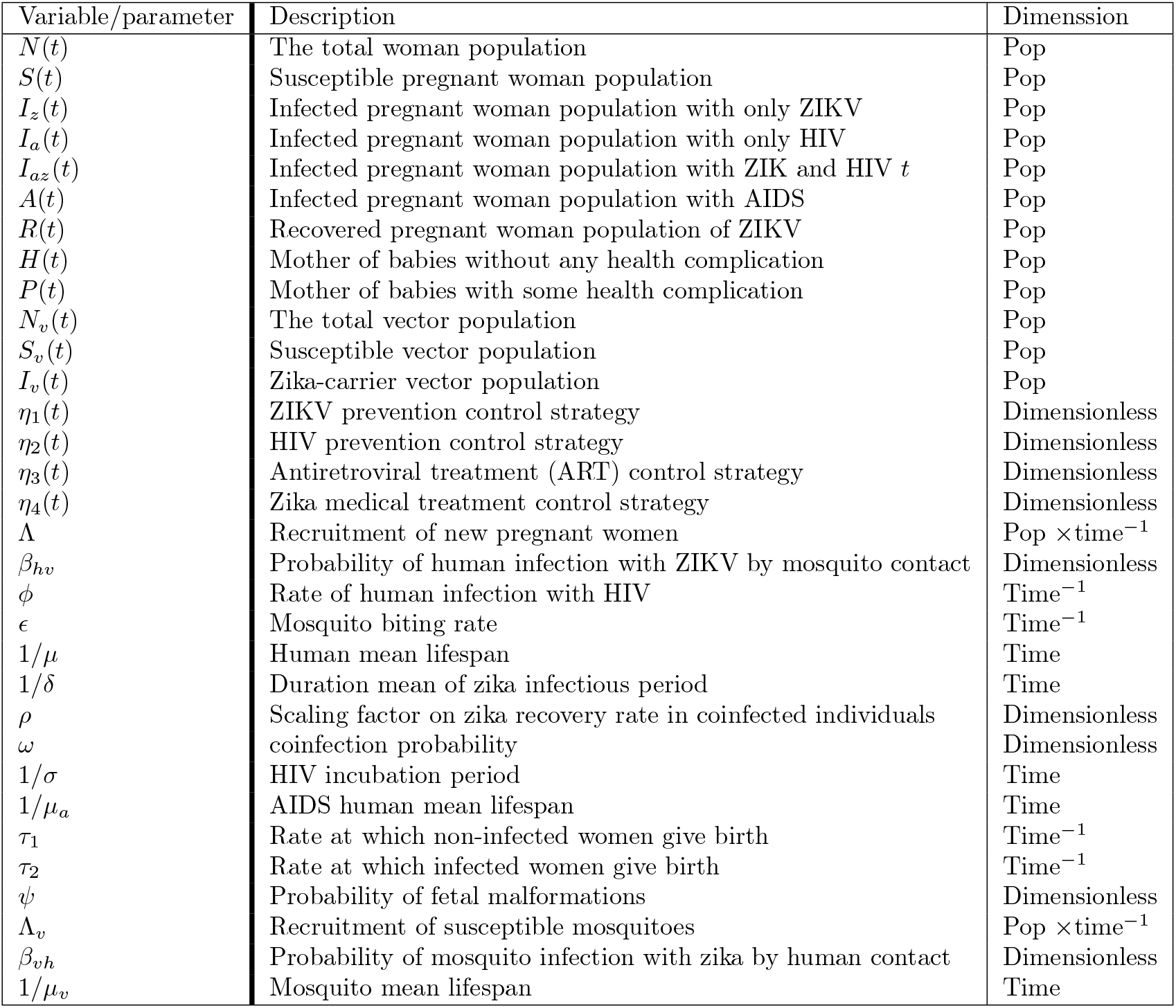
Variables and parameters incorporated in the model (1):A Description and their dimension.

**Fig 1:**
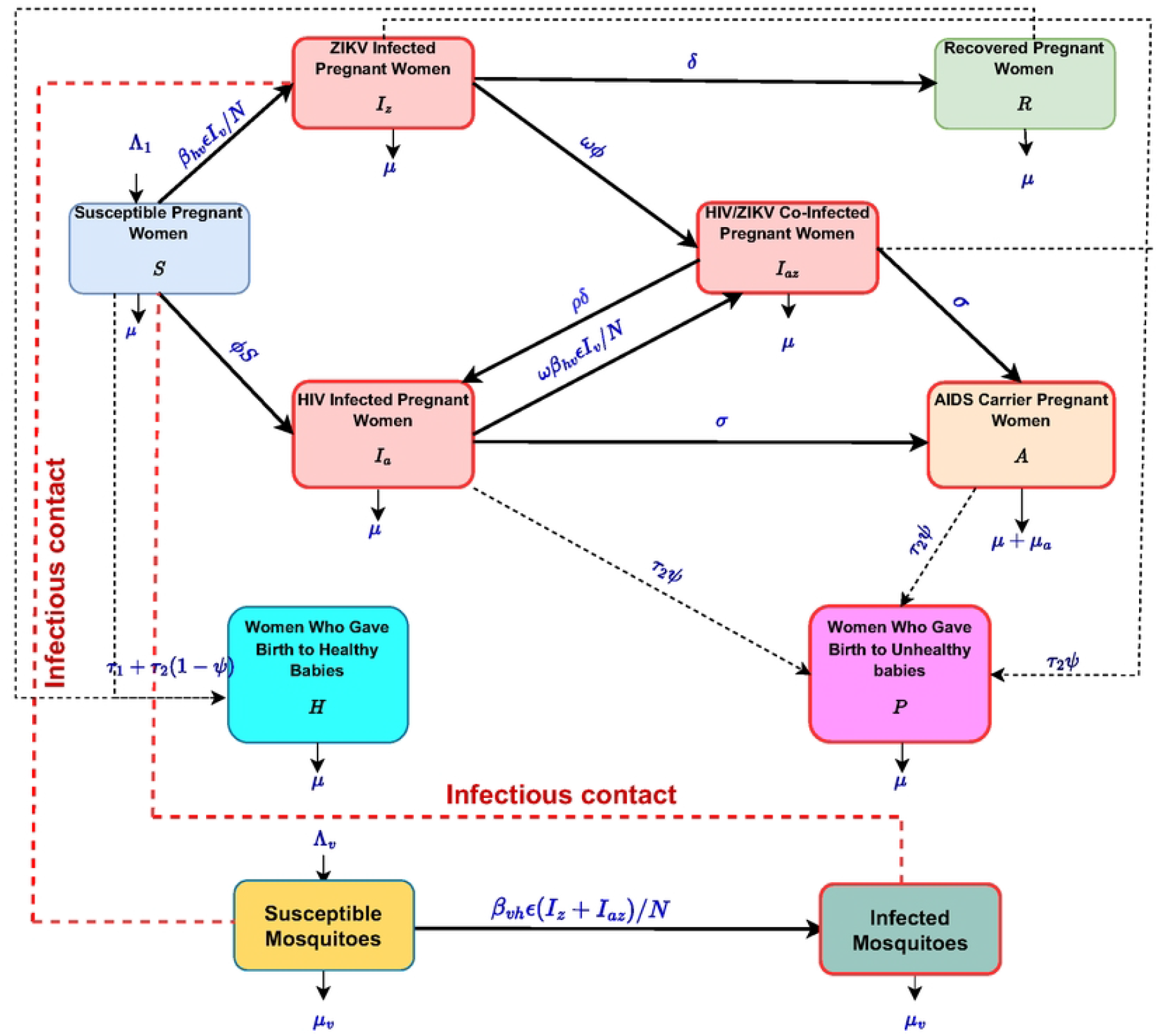
Compartmental model of the HIV/ZIKV coinfection model in pregnant women defined in (1).

**Fig 2:**
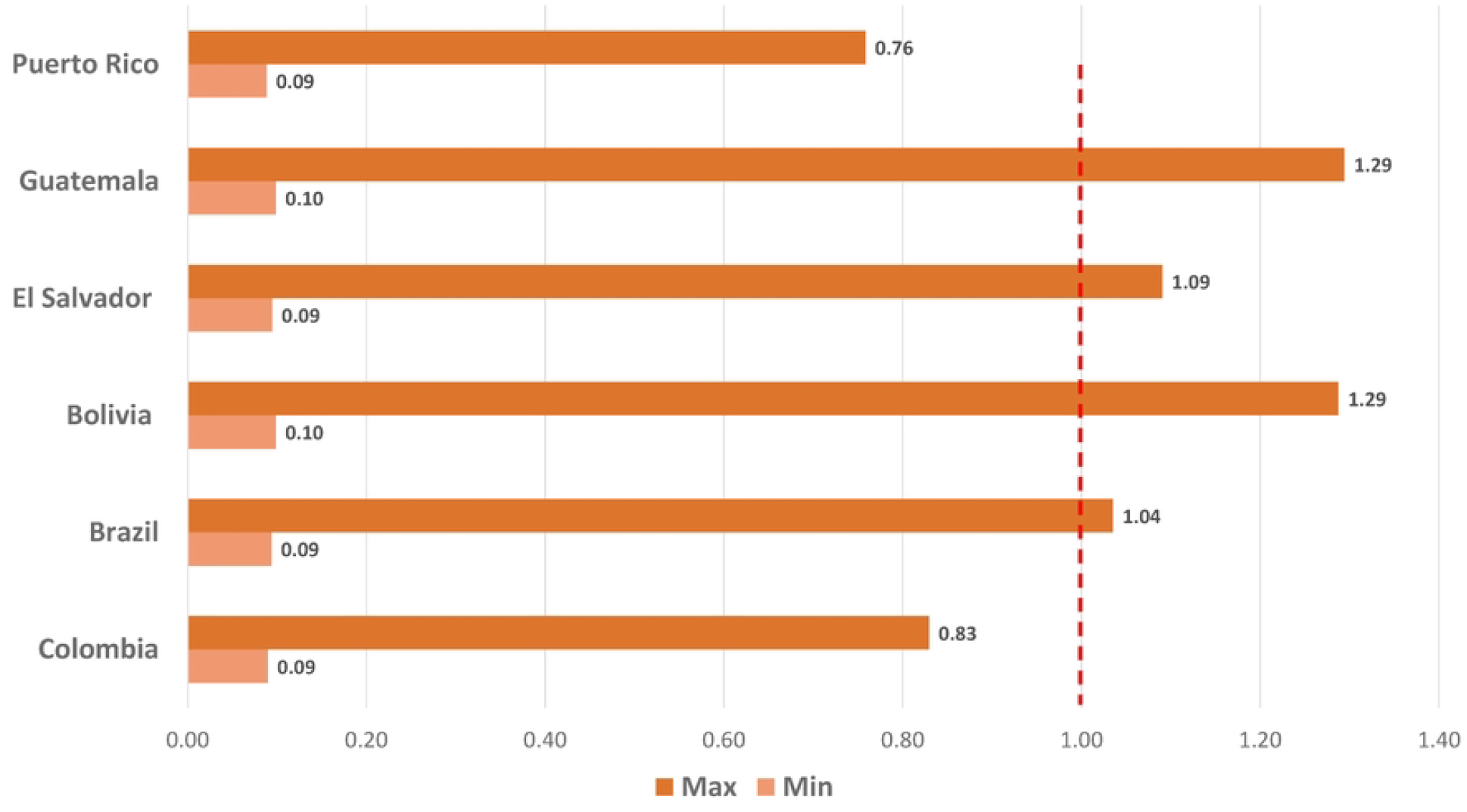
Basic reproduction number for HIV and ZIKV coinfection among pregnant women.

The transitions between these compartments are governed by a set of parameters that capture the transmission behavior of HIV and ZIKV. The rates at which non-infected (*τ*_1_) and infected pregnant women (*τ*_2_) give birth have a significant impact on the health status depicted in the compartments of women who give birth. The set of equations is available in the Appendix 6.

### 2.3 Data collection and model parameters

Our data comes from national health databases, such as the Pan American Health Organization (PAHO) reports, United Nations Programme on HIV/AIDS (UNAIDS) data, and literature reviews that highlight HIV and ZIKV providing historical and recent data on infection rates and transmission dynamics [13, 16, 22]. The primary selection criteria were the robustness of the data collection methods and the relevance to the study period (2015-2023). We chose to include Brazil, Colombia, Bolivia, El Salvador, Guatemala and Puerto Rico based on their high prevalence of HIV and ZIKV infections and their historical significance in the spread of these viruses, as reported by PAHO zika reports. Moreover, accuracy and relevance of our model are ensured by parameter selection and their values. Those are based on current epidemiological data and existing literature (Tables 2 and 3) [13, 20]. The development of mathematical methodology consisted of five stages. We started using the uncontrolled mathematical model. For this model, we calculated the basic reproductive number, performed a sensitivity analysis of parameters, and generated predictions for the compartments of women who give birth to healthy babies (*H*), and those who give birth to unhealthy babies (*P*) using different values of *τ*_1_ and *τ*_2_ and different initial population sizes of HIV and ZIKV infected pregnant women in 2022.

**Table 2:**
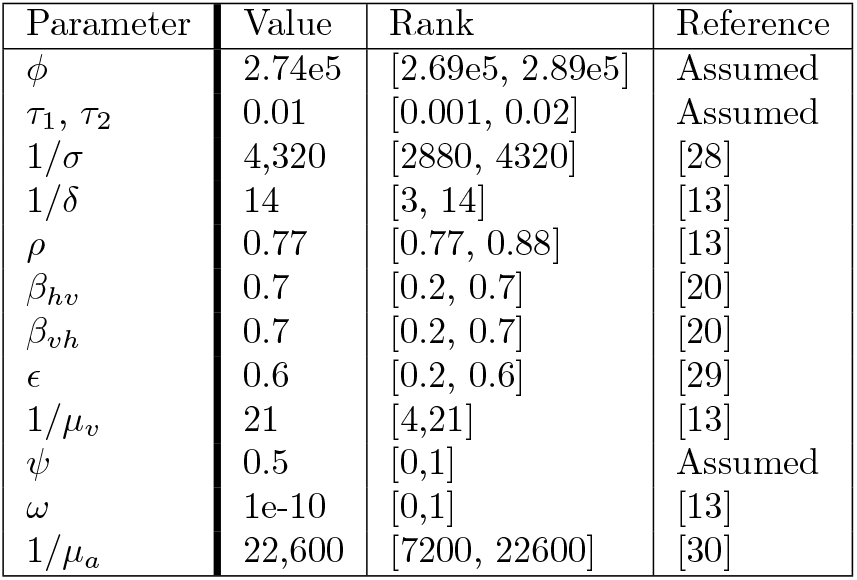
Model (1) common parameter values (Colombia, Brazil, Bolivia, El Salvador, Guatemala, Puerto Rico). Duration in days.

| Parameter | Value | Rank | Reference |
| --- | --- | --- | --- |
| $\phi$ | 2.74e5 | [2.69e5, 2.89e5] | Assumed |
| $\tau_1, \tau_2$ | 0.01 | [0.001, 0.02] | Assumed |
| $1/\sigma$ | 4,320 | [2880, 4320] | [28] |
| $1/\delta$ | 14 | [3, 14] | [13] |
| $\rho$ | 0.77 | [0.77, 0.88] | [13] |
| $\beta_{hv}$ | 0.7 | [0.2, 0.7] | [20] |
| $\beta_{vh}$ | 0.7 | [0.2, 0.7] | [20] |
| $\epsilon$ | 0.6 | [0.2, 0.6] | [29] |
| $1/\mu_v$ | 21 | [4,21] | [13] |
| $\psi$ | 0.5 | [0,1] | Assumed |
| $\omega$ | 1e-10 | [0,1] | [13] |
| $1/\mu_a$ | 22,600 | [7200, 22600] | [30] |

**Table 3:**
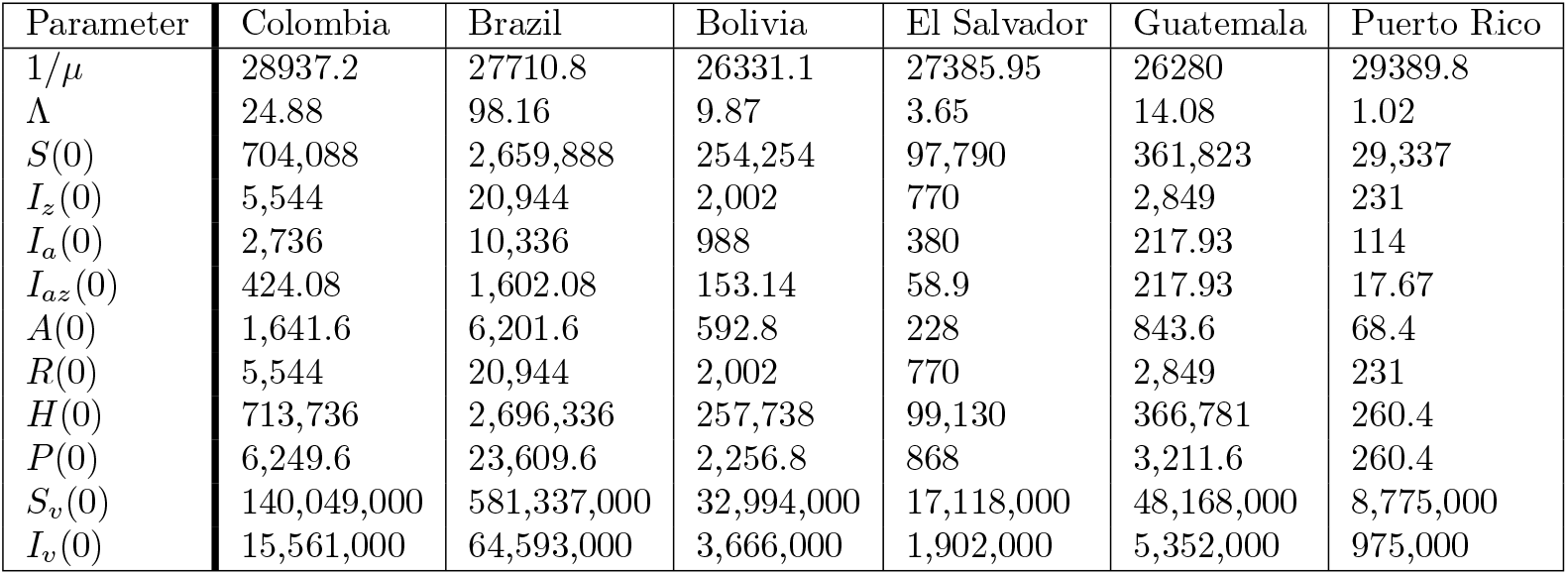
Values of no common parameters and initial conditions (2022-population size) in the model (1). Duration in days.

| Parameter | Colombia | Brazil | Bolivia | El Salvador | Guatemala | Puerto Rico |
| --- | --- | --- | --- | --- | --- | --- |
| $1/\mu$ | 28937.2 | 27710.8 | 26331.1 | 27385.95 | 26280 | 29389.8 |
| $\Lambda$ | 24.88 | 98.16 | 9.87 | 3.65 | 14.08 | 1.02 |
| $S(0)$ | 704,088 | 2,659,888 | 254,254 | 97,790 | 361,823 | 29,337 |
| $I_z(0)$ | 5,544 | 20,944 | 2,002 | 770 | 2,849 | 231 |
| $I_a(0)$ | 2,736 | 10,336 | 988 | 380 | 217.93 | 114 |
| $I_{az}(0)$ | 424.08 | 1,602.08 | 153.14 | 58.9 | 217.93 | 17.67 |
| $A(0)$ | 1,641.6 | 6,201.6 | 592.8 | 228 | 843.6 | 68.4 |
| $R(0)$ | 5,544 | 20,944 | 2,002 | 770 | 2,849 | 231 |
| $H(0)$ | 713,736 | 2,696,336 | 257,738 | 99,130 | 366,781 | 260.4 |
| $P(0)$ | 6,249.6 | 23,609.6 | 2,256.8 | 868 | 3,211.6 | 260.4 |
| $S_v(0)$ | 140,049,000 | 581,337,000 | 32,994,000 | 17,118,000 | 48,168,000 | 8,775,000 |
| $I_v(0)$ | 15,561,000 | 64,593,000 | 3,666,000 | 1,902,000 | 5,352,000 | 975,000 |

### 2.4 Analysis and simulation

The mathematical methodology development consisted of five stages. We started using the uncontrolled mathematical model. For this model, we calculated the basic reproductive number, performed a sensitivity analysis of parameters, and generated predictions for the compartments of women who give birth to healthy babies (*H*) and those who give birth to unhealthy babies (*P*) using different values of *τ*_1_ and *τ*_2_ and different initial population sizes of HIV- and ZIKV-infected pregnant women in 2022. After evaluating the sensitivity indices and understanding the parameters to which ℛ_0_ is most sensitive, we advanced to the controlled mathematical model, followed by generating detailed predictions regarding the number of pregnant women giving birth to newborns with related health issues.

The following detailed items present a step-by-step explanation of the methodology of each stage:

- **Step 1**: In a naive population regarding a specific disease, the number of persons getting infected from a single sick individual is estimated by the basic reproduction number ℛ_0_. It provides insight into how rapidly an infection might spread, which in turn informs the types of actions that might be necessary to control it. We initially calculated the ℛ_0_ for HIV and ZIKV separately, then examined what happens when they co-occur in the same model. The Next Generation Method [23] was used in order to determine the ℛ_0_.
- **Step 2**: We wanted to know which parameter has the most influence on how quickly HIV and ZIKV can spread among pregnant women, so we used a local sensitivity analysis [24]. A sensitivity index tells us how much ℛ_0_ changes when a particular parameter varies. If the sensitivity index is positive for a specific parameter, it means that increasing that parameter will also increase ℛ_0_ and that, thus, the transmission of the disease is higher. However, if the sensitivity index is negative, it means that increasing that parameter will decrease ℛ_0_. We focus on the effect of one parameter at a time in that method, which involves a mathematical process called partial derivative, which represents ℛ_0_’s rating change. Multiplying it by the parameter value divided by ℛ_0_ allows us to obtain the sensitivity index.
- **Step 3**: We studied the influence of the variation of the model’s parameter on the number of women giving birth to healthy and unhealthy babies. We realized simulations for three different ranges (low, medium, and high) of values of the rates of women giving birth, denoted as 1 and 2. Additionally, we performed simulations with different initial conditions of pregnant women infected with either HIV or ZIKV to see how newborns’ health would be impacted.
- **Step 4**: Four control variables were then incorporated into our model: measures to reduce mosquito bites (*η*_1_), strategies to prevent sexual transmission of HIV (*eta*_2_), the use of ART to reduce the progression of HIV to AIDS (*η*_3_), and medical treatment of ZIKV (*η*_4_). These control measures were applied over a specific period ([0, *T*]) and ranged in effectiveness from 0, meaning no control over the virus transmission, to 1, synonymous with full control. We then used Pontryagin’s Maximum Principle to find the best combination of these controls to minimize the number of infections and the associated costs [25]. The detailed mathematical formulation and proofs are provided in Appendix 6.
- **Step 5**: We used the backward-forward sweep method with the ODE45 function in MATLAB, which helps with accurate numerical integration during each iteration [26, 27]. The algorithm implemented in this method is described as follows:
  - Initial guess of the control measures value.
  - Equation solving for the state variables, forward in time, such as the number of infected individuals.
  - Equation solving for the adjoint variables backward in time for optimization purposes.
  - Update of the control measures based on these solutions.

This process was then repeated until the control measures were stabilized to ensure the accuracy of the results. These simulations aimed to find the optimal combination of control measures. A detailed explanation of the optimal control analysis and the underlying mathematics can be found in Appendix 6.

## 3 Results

### 3.1 Basic reproduction number and predicted health outcomes in newborns

#### 3.1.1 Basic reproduction number ℛ_0_

In a population of susceptible individuals, the number of secondary infections from an infected person is given by the basic reproduction number ℛ_0_. In this study, we calculated ℛ_0_ for both HIV and ZIKV coinfection using the Next Generation Matrix approach. The detailed calculations for ℛ_0_ can be found in Appendix 6 and Appendix **??**.

When ℛ_0_ exceeds one, there is a higher transmission of the disease since one single infected person affects many people and thus a risk of an epidemic. Conversely, implementing public health measures aims to reduce ℛ_0_ below one, which means controlled transmission levels and potentially the extinction of the disease.

The ℛ_0_ associated with both viruses’ minimum was 0.09 in all countries of interest, except Bolivia and Guatemala, where it was estimated at 0.1. However, maximum values ℛ_0_e observed to be 0.76 in Puerto Rico, 0.83 in Colombia, 1.04 in Brazil, 1.09 in El Salvador, and 1.29 in Guatemala and Bolivia. Even if those values are close to 1, they could still lead to a rise of altered births in the appropriate conditions. Application of control measures to limit the coinfection and reduce related health complications in newborns is crucial.

#### 3.1.2 Sensitivity analysis of key parameters

The basic reproduction number ℛ_0_ is impacted by the parameters of the model. We performed a sensitivity analysis for the basic reproduction number of both HIV and ZIKV combined to identify which parameters it is the most sensitive to. The goal is to indicate the intervention strategies to prioritize to effectively reduce ℛ_0_. The detailed results for each country and each pathology are provided in Appendix 6.

The parameters having the biggest positive influence on the ℛ_0_ were estimated to be the human death rate (*µ*) as well as the mosquito biting rate (*ϵ*). This means that when their values increase, the basic reproduction associated with the coinfection increases as well. To a lesser extent, the rate at which non-infected women give birth (*τ*_1_), the probabilities of human infection with ZIKV by mosquito contact (*β*_*hv*_) and the probability of mosquito infection with ZIKV by human contact (*β*_*vh*_) as well as the recruitment of susceptible mosquitoes (Λ_*v*_), also are positively associated with ℛ_0_. On the other hand, an increase in the recruitment of new pregnant women (Λ) or a decrease in the mean duration of the zika infectious period will lead to a decrease in the value in the ℛ_0_. All the other parameters included in our model had a sensitivity index of 0.

#### 3.1.3 Model predictions for newborn health outcomes

We used the model to obtain predictions about the number of women who give birth to babies with health complications due to HIV and ZIKV maternal coinfection using the model’s equations. As expected, an increase in this population over time is expected when no intervention measures are applied to the model.

A table with the accumulated data regarding our different variables of interest for the 2022-2025 period can be found in Appendix 6. Our model predictions indicate that the mean percentage of women giving birth to unhealthy babies in the countries studied during that period is 13.9%. Among them, Brazil exhibited the highest proportion of women giving birth to unhealthy births, with 39%, whereas Guatemala had the lowest at 5%. Colombia and El Salvador both showed approximately 6.5% of related altered births, while Puerto Rico had a higher rate with 18.5%.

### 3.2 Impact of parameters and population sizes in health outcomes

#### 3.2.1 Initial infection levels

The initial conditions for the number of infected pregnant women with both HIV and ZIKV will impact newborns’ health outcomes. We performed simulations to determine the number of women giving birth to unhealthy babies for varying levels of infections.

Figures 3 and 4 represent the percentage increase in unhealthy births for different levels of ZIKV *I*_*z*_(0) and HIV *I*_*a*_(0) circulation among pregnant women over 1,000 days. For ZIKV, Figure 3 shows that the simple introduction of the virus among pregnant women, when *I*_*z*_(0) shifts from 0% to 0.77%, causes a surge in the number of women who give birth to unhealthy babies, with a minimum of 436% in Guatemala and a maximum of 1,022% in Puerto Rico. This pronounced impact is due to the high transmission potential of ZIKV during pregnancy 8. Interestingly, further increase in ZIKV circulation among pregnant women does not have a significant effect on newborn’s health, with a mean increase of 1.38% among all studied countries when ZIKV levels goes from 0.77% to 23.43% 8.

**Fig 3:**
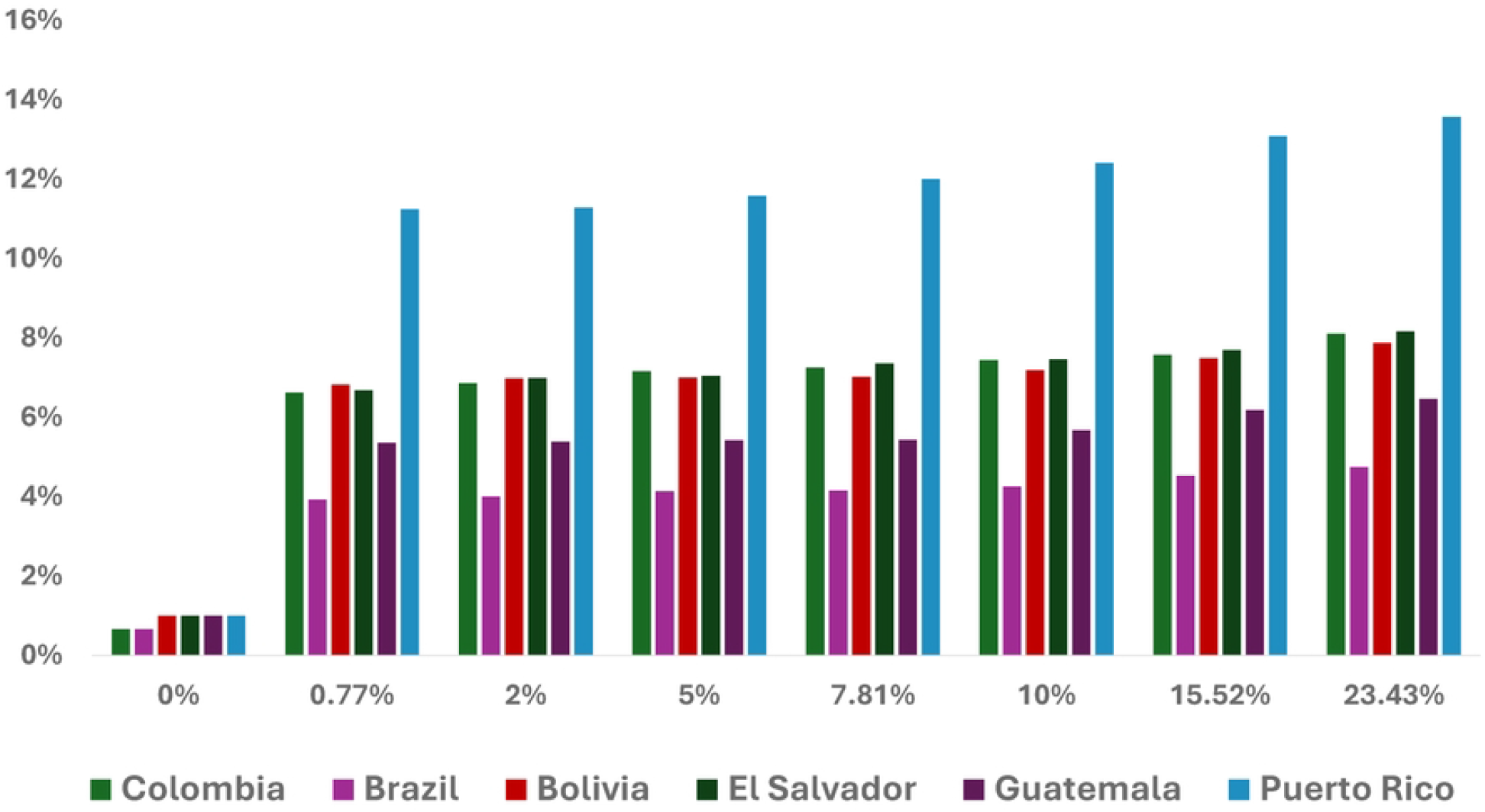
Percentage increase in related unhealthy births for different ZIKV levels among pregnant women.

**Fig 4:**
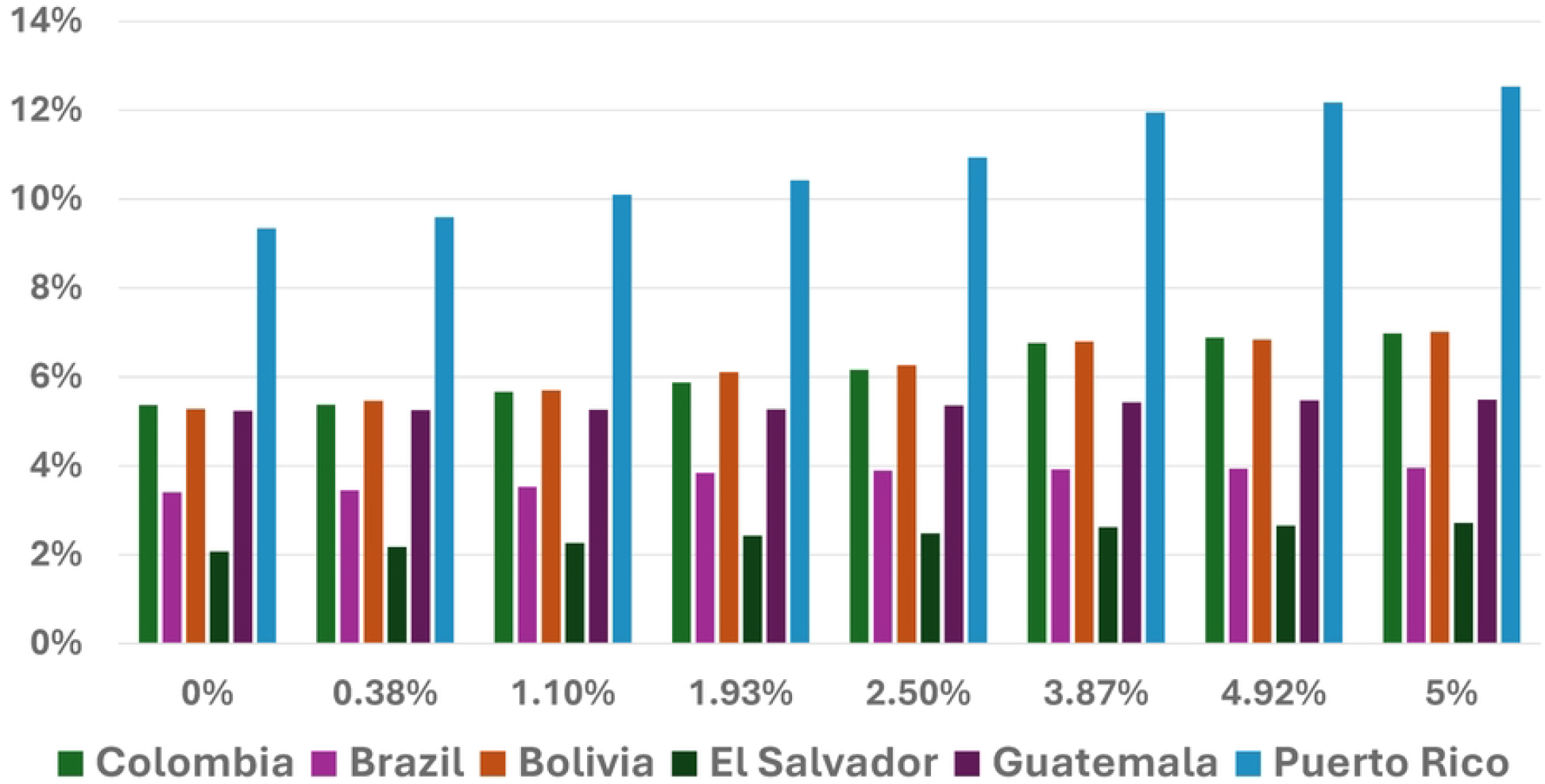
Percentage increase in related unhealthy births for different HIV levels among pregnant women.

In contrast, Figure 4 shows that the impact of the initial number of HIV-infected pregnant women (*I*_*a*_(0)) leads to a more subtle increase in the number of unhealthy births, though at a lower rate. All the countries follow a similar trend, with Puerto Rico the most impacted, when *I*_*a*_(0) goes from 0% to 5%, there is a 3.19% increase in altered births. This country also has the highest overall percentage of unhealthy births 6. Conversely, Guatemala is the least impacted country with only a 0.24% increase when HIV infection levels among pregnant women rise from 0% to 5% 6.

### 3.3 Evaluation and Comparison of Control Strategies

When our model doesn’t take into account the implementation of any control measure, it reveals an increase in the incidence of health complications related to maternal infection with either HIV or ZIKV across all the studied countries when the virus prevalence increases. However, acknowledging the real-world response to the pandemic, governments have implemented interventions to mitigate zika infections in their populations. We thus used sensitivity indices results and our understanding of the virus dynamics to incorporate four intervention strategies into our model and assess their relative efficiency and effects on newborn health.

Figure 5 illustrates the dynamics of the activation of the different HIV and ZIKV strategies over 1,000 days to control the number of unhealthy births in each country optimally, with 0 being synonymous with no control and 1 total control. Medical treatment of ZIKV was the measure that needed the least activation over time in all the studied countries. On the other hand, the use of antiretroviral drugs to treat HIV infection among pregnant women is the measure that was the most activated in Bolivia, Colombia, El Salvador, and Puerto Rico in order to limit altered consequences on neonatal health. HIV and ZIKV prevention were also necessary in every country in order to optimally control the coinfection among pregnant women. In our modelling approach, control measures were triggered only when an increase in zika cases was observed. Regional variability in demographic factors, such as the number of infected pregnant women or the total population, is in part responsible for the fluctuation of control measures’ effectiveness between countries.

**Fig 5:**
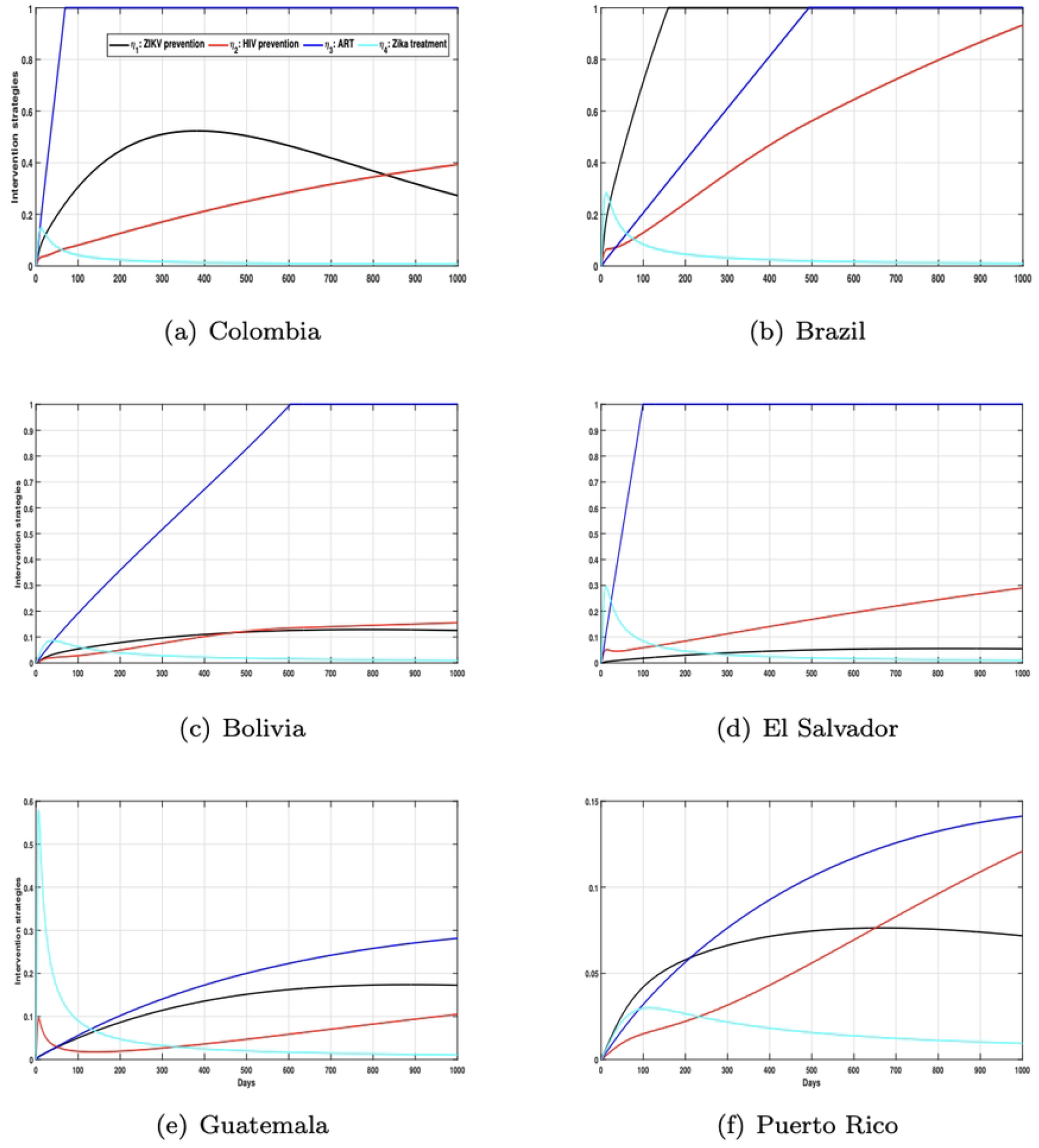
Simulations of the implementation of control strategies over time. In this context, *η*_1_ represents zika prevention, *η*_2_ denotes HIV prevention, *η*_3_ represents antiretroviral therapy, and *η*_4_ stands for medical treatment for zika.

## 4 Discussion

Our primary aim was to improve our understanding of the dynamics around the HIV-ZIKV coinfection among pregnant women. Its effects on their newborns’ health considering different epidemiological factors were also part of our primary concern. The mathematical model developed accounted for vectorial transmission of ZIKV as well as the likelihood of coinfection with both viruses and intervention strategies, such as antiretroviral treatment for HIV-positive individuals, measures to prevent zika and HIV, and medical treatment for zika.

To validate our mathematical model, we used data from a Brazilian cohort of pregnant women infected with HIV during the 2015 zika epidemic [15]. We extrapolated these data to other countries, including Colombia, El Salvador, Guatemala, Puerto Rico, and Bolivia. Hence, our study pioneers the examination of the distinctive context of maternal ZIKV infection in HIV-seropositive pregnant women, a subject that has not been thoroughly investigated since only two other models for HIV/ZIKV coinfection have been developed as far as we are aware (see e.g., [13, 14]). Nonetheless, the models were validated using data from Colombia and Brazil. Most importantly, neither of them integrated characteristics to address the effects of such a coinfection on the health of babies, nor were they intended for the community of pregnant women.

Importantly, our simulations demonstrated that even a slight increase in ZIKV infections among pregnant women could have a disproportionately negative impact on their newborns health. This finding highlights the necessity for targeted interventions to limit ZIKV transmission, especially among this vulnerable population. Including control measures in our model revealed that combining long-term public health measures, such as HIV or ZIKV prevention, would be the most effective strategy to reduce levels of ZIKV and HIV infection among pregnant women and therefore improve newborn health outcomes.

We estimated the range of the basic reproduction number for our coinfection mathematical model. In general, the maximum estimated values were close to or slightly above 1 in the studied countries. Those results suggest a moderate potential for an epidemic under studied conditions and align with previous studies in some regions but differ in others. For instance, a study conducted during the 2015 zika epidemic in Rio de Janeiro estimated an ℛ_0_ for ZIKV of 2.33 [31] significantly higher than ours. Moreover, a model reported an average ℛ_0_ of 4.3 for ZIKV in El Salvador [32]. The higher ℛ_0_ values from these studies could be caused by differences in demographic dynamics or specific environmental conditions that we may not have accounted for. Regarding the ℛ_0_, we found a minimum of 0.09 and a maximum of 1.29 across our countries of interest, with the highest values for Guatemala and Bolivia. These results are lower than the ones from a study that reported a minimumℛ_0_ related to HIV of approximately 0.4719 and of 0.23606 for the ℛ_0_ related to ZIKV in the general population of Brazil and Colombia [13]. Our focus on the pregnant women’s population might impact the ℛ_0_ values obtained and could explain why they are smaller than the ones from studies regarding the general population. However, it is necessary to pursue public health efforts in this high-risk region to keep ℛ_0_ below 1 to prevent outbreaks. We mentioned that rising levels of ZIKV infection among pregnant women had a more significant negative impact on newborn health compared to HIV. Consistent with previous studies, our results showed an increase in congenital abnormalities associated with ZIKV maternal infections. For example, a study conducted among HIV-seropositive pregnant women in Rio (Brazil) reported that ZIKV infection among their cohort of pregnant women was the primary factor associated with neurological malformations in infants [15]. Those results are consistent with our findings of a 0.14% incidence of central nervous system abnormalities among newborns in Brazil at the highest levels of pregnant women’s HIV infection. However, our study differed by including a broader range of health outcomes, potentially explaining variations in the incidence rates. Moreover, between 110 and 270 babies were born with microcephaly out of 10,300 ZIKV-positive-pregnancies, as reported by a study conducted in Puerto Rico in 2016 [33] while our model estimated 129 as the number of pregnant women giving birth to unhealthy babies in the country between 2022 and 2025 6.

Our model, which predicted varying levels of newborn health complications across countries, supports the idea that, even if it generally poses a severe risk to neonatal health, the impact of ZIKV can vary significantly based on local factors. Our findings converge with the literature, where intervention strategies addressing both sexual transmission and mosquito control seem to be the most effective way to control both viruses [14]. Our sensitivity analysis results underscored that coinfection transmission is mostly affected by the mosquito biting rate and the overall population of ZIKV vectors. Moreover, control of ZIKV is complex since studies highlighted the mathematical phenomenon of backward bifurcation observed in the context of the ZIKV epidemic, where keeping the ℛ_0_ below 1 is necessary but may not be sufficient to eliminate the disease [34],emphasizing the need for strong and sustained public health interventions and surveillance, especially in regions with high mosquito density and limited healthcare access.

Different factors could explain the differences between our results and those from other studies. Firstly, the basic assumptions of the models vary between models. For example, we assumed to have a constant vector population as well as a homogeneous population mixing. Additionally, the lower ℛ_0_ and the different health impacts observed compared to other studies might be explained by the fact that we focused on the population of pregnant women, which requires specific healthcare needs. Regional and environmental factors impacting competent ZIKV vectors may also contribute to variations among results. Finally, we included a broad spectrum of health outcomes for newborns of infected pregnant women, leading to different conclusions compared to studies focused on specific conditions like microcephaly.

Our results are thus consistent with the existing literature highlighting the greater effect of ZIKV compared to HIV on newborn health in the context of maternal coinfection. However, different studies report different ranges of basic reproduction numbers for ZIKV, demonstrating the importance of using adequate parameter values to improve modelling results. Precise results would help decision-makers in their choice of control measures.

### 4.1 Study limitation

Our results regarding the dynamics of HIV and ZIKV coinfection among pregnant women and its effects on newborn health could be impacted by several potential biases, which would lead to a lack of precision. First, our results are observable at the population level and may not accurately represent individual-level relationships. This bias, known as ecological fallacy, could have been introduced since we used aggregated national data for population data, such as the number of pregnant women in each country. We diminished this risk by ensuring the high quality of our data by choosing reliable sources such as peer-reviewed literature and national health databases, but this limitation is inherent in many large-scale epidemiological studies and could lead to misinterpretation. We extracted the demographic data from the population division of the United Nations data portal [35]. This allowed us to obtain evidence that will contribute to ensuring the robustness and consistency of our study, since it is true that our model’s accuracy primarily depends on the quality and precision of the input parameters’ values, varying across countries. Differences in health data availability could thus introduce information bias. Sensitivity analysis, which allows us to discover the effects of key parameters on the model outcomes, contributes to addressing this issue. We also used biologically credible ranges for the parameter values extracted from the literature to capture potential variability. Intrinsic uncertainty in parameter estimation cannot be eliminated, meaning that our results should be interpreted with caution.

Thirdly, we assumed homogeneous mixing within populations for our model. This means that every individual has the same likelihood of coming into contact with any other one. We thus oversimplify complex interactions not only between but also within populations, not accounting for social behaviors or heterogeneity in healthcare access, for example. Adjustment of population data to reflect regional demographics and variations was attempted to manage this effect. Moreover, uniform application of the model to all the countries of study, primarily based on population size, introduces further limitations. This approach allows for results comparison and analysis across the six countries of our study. Nonetheless, there are significant local variations in HIV and ZIKV viral transmission dynamics, particularly those influenced by cultural, environmental, or socio-economic factors. For instance, to obtain values of some of our parameters, we used data from the Brazilian comprehensive surveillance system during the 2015 ZIKV outbreak, which provided detailed data regarding viral transmission and congenital defects among newborns. These were not available for some of the other countries of interest. This potentially leads to discrepancies in our model accuracy across regions. Additionally, climatic factors, such as temperature or rainfall levels, are critical determinants of mosquito populations and thus ZIKV transmission. However, we did not incorporate those factors in our model, which alters our ability to reflect real-world scenarios. Furthermore, our results did not account for the potential development of population immunity against ZIKV. This could cause an alteration of infection levels over time in regions with previous viral exposure. This omission is particularly relevant given the asymptomatic nature of ZIKV in approximately 80% of cases; this omission is particularly relevant since it complicates the accuracy and detection of immunity within populations. Serological testing and polymerase chain reaction (PCR) are the main detection methods for ZIKV, but are highly dependent on the timing of testing. Cross-reactivity or prior exposure to other arboviruses can lead to misinterpretation [36]. It is thus complicated to obtain an accurate estimation of ZIKV prevalence and assess control strategies efficiency. Overall, our model allows for a better understanding of the potential impacts of HIV and ZIKV coinfections on maternal and neonatal health, but our results should be interpreted with caution and take into account the limitations that we pointed out. Future research should focus on incorporating more precise data to improve results accuracy. Accounting for regional heterogeneity and including climatic as well as immunological variables that could influence HIV and ZIKV dynamics would contribute to the improvement of those results.

## 5 Conclusion

Our innovative study emphasizes the effects of HIV and ZIKV coinfection among pregnant women on newborn health. It is thus crucial to prevent infection with both HIV and ZIKV in this vulnerable population to prevent a public health crisis and not have to face the same challenges as those faced during the 2015 ZIKV epidemic in Latin American and Caribbean countries. Continued surveillance and concurrent efforts from governments and public health institutions are the keys to reducing the burden of this coinfection among pregnant women. In the end, mathematical modelling proves to be a useful tool in public health, helping with decision-making processes and offering estimates for distinct scenarios, knowing that projections show that ZIKV is becoming more common in endemic nations and may eventually spread to other areas, including North America.

## Data Availability

This study is based exclusively on secondary data obtained from previously published studies and publicly available sources. All relevant data are included within the manuscript and its references. No new data were generated or analyzed for this study.

## 6 Supporting information

### S1 Appendix. The mathematical model formulation

Under the assumptions considered in Section 2.2, we have the following mathematical model:

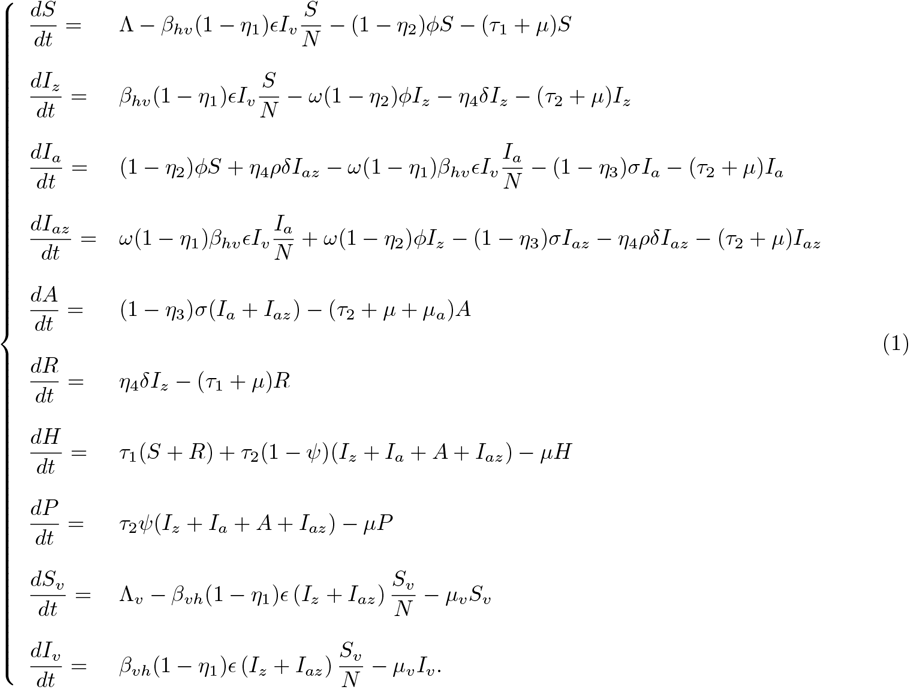

For the above model, without controls, we state the following biological interest region

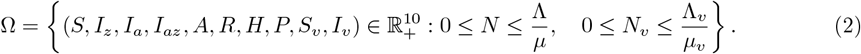

Therefore, it is enough to consider the dynamics of (1) in Ω.

### S2 Appendix. The optimal control problem analysis

One of our objectives will be to minimize the cost associated with the following objective functional:

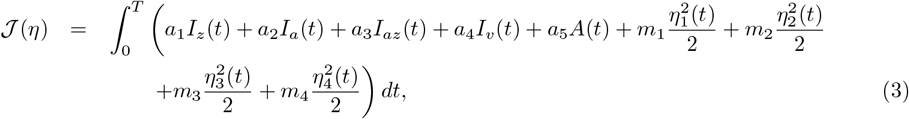

with *η* = (*η*_1_(*t*), *η*_2_(*t*), *η*_3_(*t*), *η*_*a*_(*t*)), and *a*_1_, *a*_2_, *a*_3_, *a*_4_, *a*_5_, *m*_1_, *m*_2_, *m*_3_ and *m*_4_ are positive weights (see Table 1). We also state the following boundary conditions:

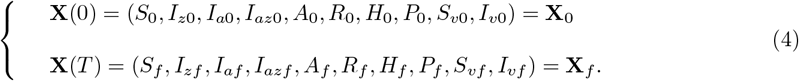

In this context, we examine an optimal control problem (OCP) consisting of the equations (1), (3) and (4). Hence, our aim is to identify an optimal control *η*^∗^(*t*) determined as

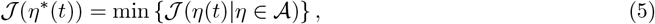

with a set of controls 𝒜 defined as

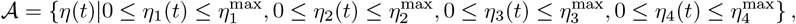

where 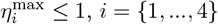 and *η* is Lebesgue measurable. In order to define the formulation of our OCP using Pontryagin’s Maximum Principle (PMP) [37], we have the Lagrangian as

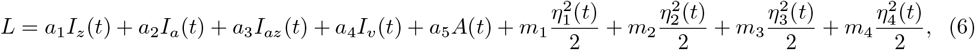

and we determine the Hamiltonian function as

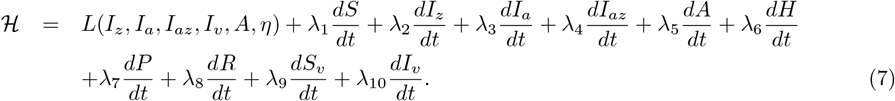

In the remainder, we investigate the minimum value of Lagrangian (6). Firstly, we must prove the existence of the optimal control *η*^∗^ according to the controlled system (1).

#### Proposition 6.1.

*There exists an optimal control η*^∗^ *such that*

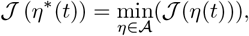

*subject to the controlled system* (1) *with initial conditions as* **X**_**0**_.

In the following, we apply PMP [37] to provide a characterization of an optimal control solution to the Hamiltonian (7) subject to the OCP (1). If (*X*^∗^, *η*^∗^) is an optimal solution for the controlled system (1), then there exists a non trivial vector function *λ* = (*λ*_1_, *λ*_2_, *λ*_3_, *λ*_4_, *λ*_5_, *λ*_6_, *λ*_7_, *λ*_8_, *λ*_9_, *λ*_10_), such that

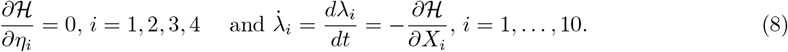

#### Proposition 6.2.

*Let* 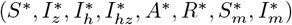 *be the optimal state variables solution associated to the optimal control variable η*^∗^ *subject to the control problem* (5). *Then, there exists an adjoint vector p that satisfies the controlled system* (1), *with transversality conditions p*_*i*_(*T*) = 0, *for i* = 1, …, 8, *where the optimal controls are*

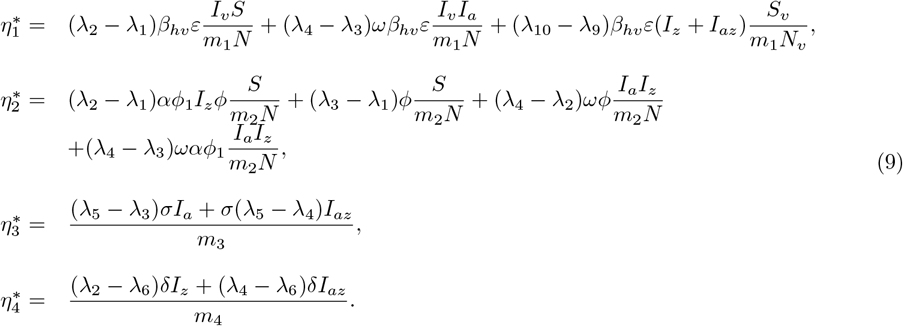

All state variables and controls are non-negative and, for *i* = {1, 2, 3, 4}, the set of control variables *η*_*i*_ ∈ 𝒜 is also convex and closed. We note that the boundedness of the optimal system (1) determines the compactness for the existence of the optimal control. Moreover, there exists a constant *ν >* 1, *ω*_1_ = min(*d*_1_, *d*_2_, *d*_3_), and *ω*_2_ *>* 0 such that

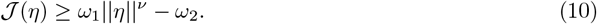

Therefore, according to [38], the controlled system (1) admits an optimal control solution *η*^∗^. We have

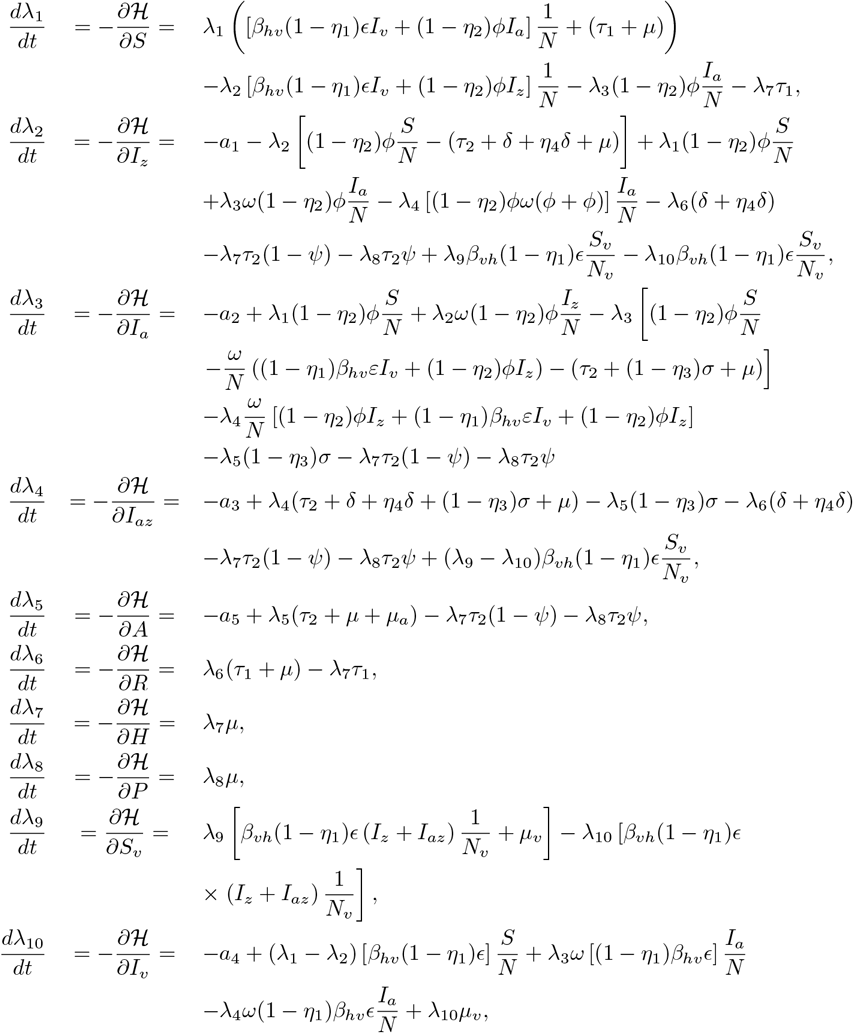

with transversality conditions *λ*_*i*_(*T*) = 0, for *i* = {1, 2, 3, 4, 5, 6, 7, 8, 9, 10}. According to PMP, the optimal conditions are

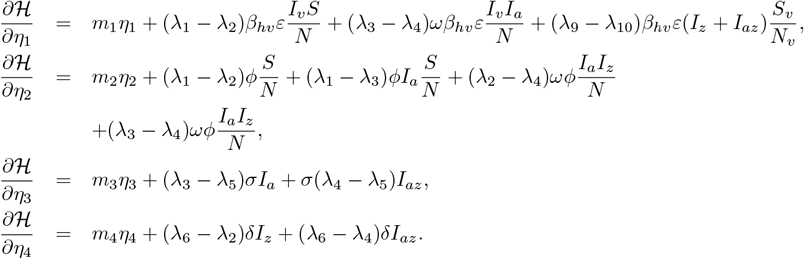

Hence, we get assertions (6.2). This completes the proof.

### S3 Appendix. The basic reproduction number and its sensitivity indices

The disease-free equilibrium (DFE) of (1) is given by setting *I*_*z*_ = *I*_*a*_ = *I*_*az*_ = *R* = *P* = *I*_*v*_ = 0. After some algebraic manipulation we get

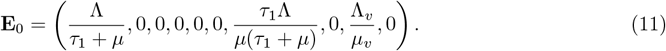

The basic reproduction number for Model (1) (ℛ_0_), can be computed using the next-generation operator [39]. Thus, the matrices **F** and **V** are given by

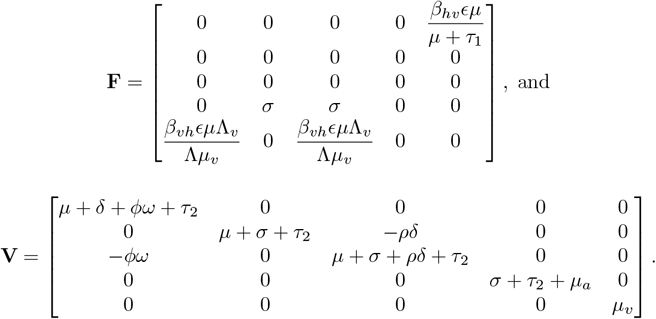

Here, the matrix **FV**^−1^ is given by

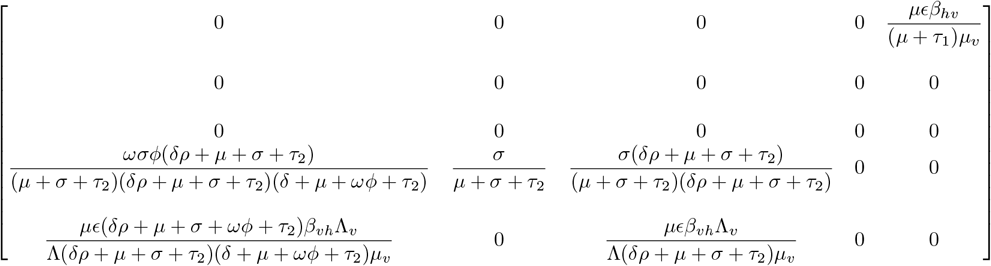

The eigenvalues of the above matrix are *λ*_1,2,3_ = 0 (three times), and

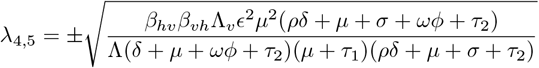

Then, the basic reproduction number for Model (1), which is defined as the spectral radius (maximum absolute eigenvalue) of the next generation matrix (**FV**^−1^) is given by:

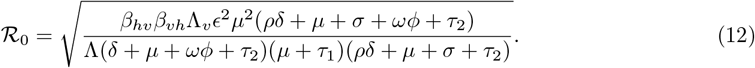

We can gain insight into the impact of parameter variations on the value of ℛ_0_ by performing a local sensitivity analysis on the model parameters. The direction of change can be determined by the sign of the sensitivity index, where a positive index for a specific parameter indicates that increasing that parameter will lead to an increase in ℛ_0_, and vice-versa.

The definition of the normalized sensitivity index for a variable with respect to a parameter is the ratio between the relative change in the variable and the relative change in the parameter [40].

Specifically, the normalized sensitivity index of the variable *X*, which is differentially dependent on the parameter *p*, can be expressed as follows:

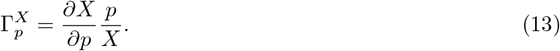

For example, to compute the sensitive index of ℛ_0_ in (12) with respect to *τ*_1_ by using the equation (13) we get

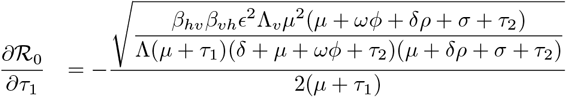

Then, it is enough to compute the expression for

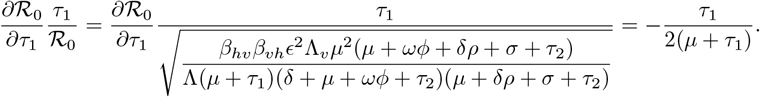

### S4 Appendix. Visual representation of the sensitivity index analysis for HIV and ZIKV basic reproduction numbers

**Fig 6:**
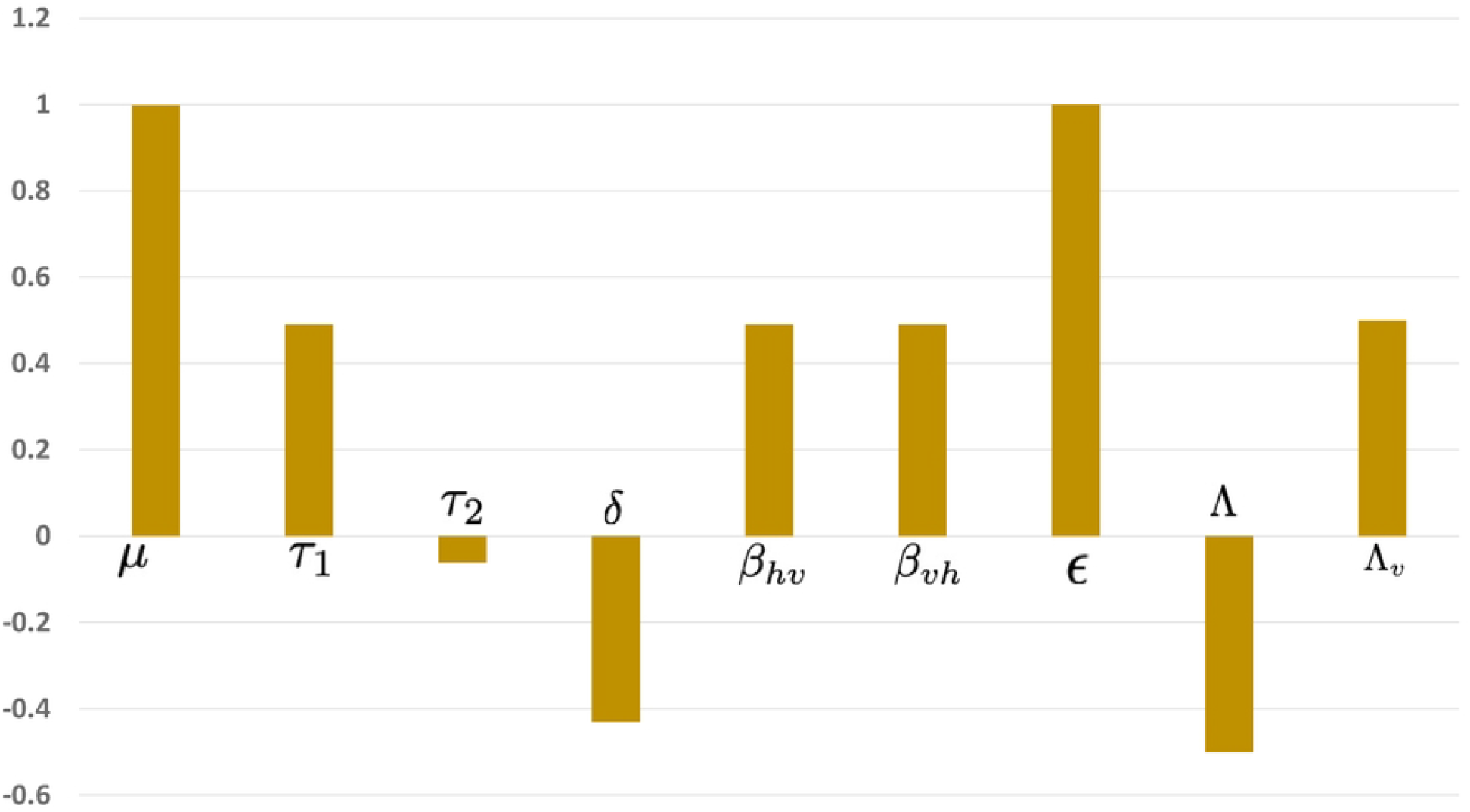
Normalized sensitivity index for the parameters {*µ, τ*_1_, *τ*_2_, *δ, β*_*hv*_, *β*_*vh*_, *ϵ*, Λ, Λ_*v*_} to ℛ_0_. Parameters involved in ℛ_0_ not included in this representation have index zero.

### S5 Appendix. Simulations of women giving birth to healthy babies for different fetal malformation probabilities. Low probability(*ψ* = 0.1**), medium probability (***ψ* = 0.5**), high probability (***ψ* = 0.9)

**Fig 7:**
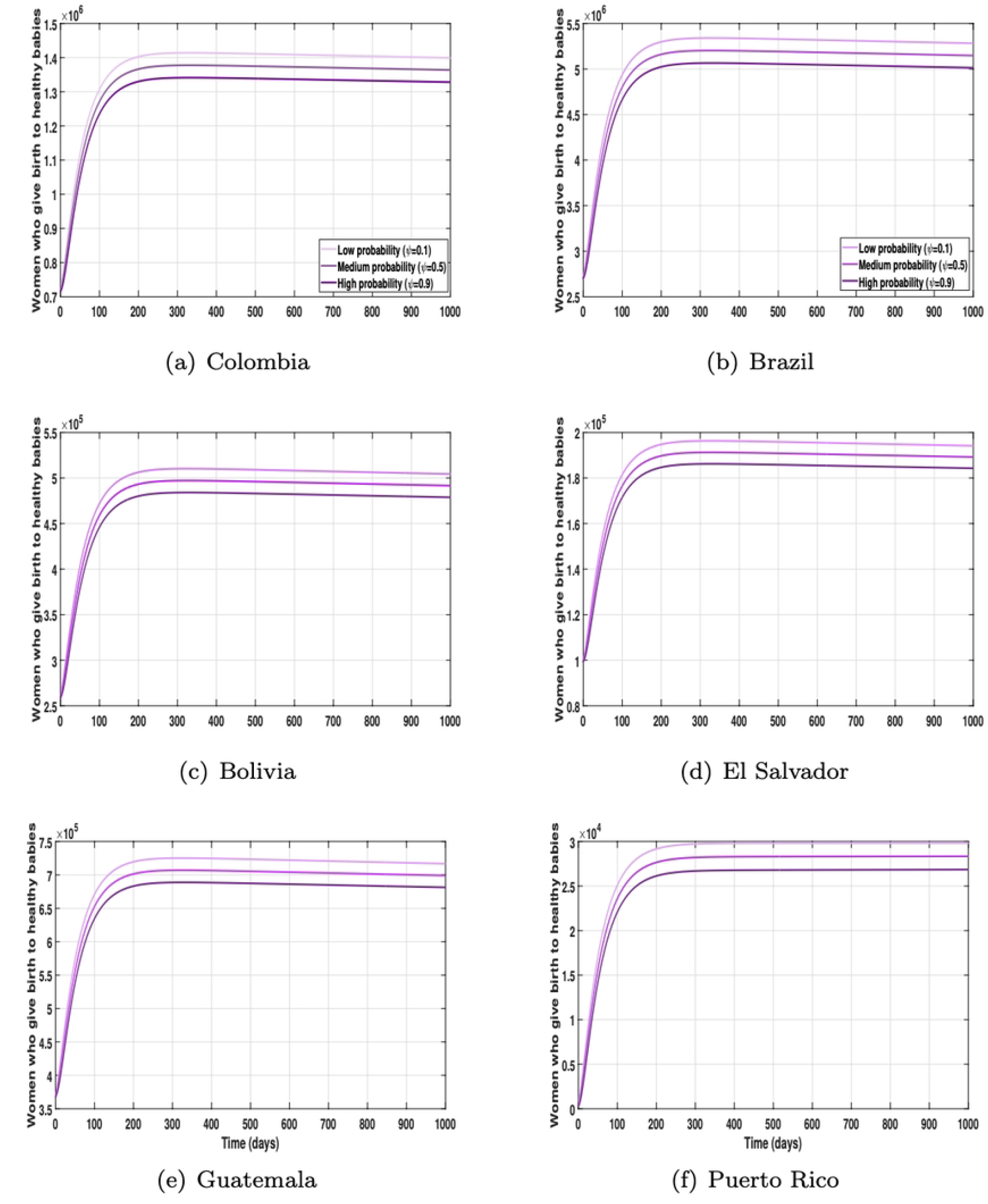
Simulations of women giving birth to healthy babies for different fetal malformation probabilities. Low probability(*ψ* = 0.1), medium probability (*ψ* = 0.5), high probability (*ψ* = 0.9).

**Table 4:** Densities of women giving birth to healthy babies for different fetal malformation probabilities over 1,000 days. Low probability(*ψ* = 0.1), medium probability (*ψ* = 0.5), high probability (*ψ* = 0.9).

| Probability | Colombia | Brazil | Bolivia | El Salvador | Guatemala | P. Rico |
| --- | --- | --- | --- | --- | --- | --- |
| Low | 1.40e6 | 5.30e6 | 5.10e5 | 1.92e5 | 7.25e5 | 3.00e4 |
| Medium | 1.37e6 | 5.20e6 | 4.97e5 | 1.90e5 | 7.00e5 | 2.81e4 |
| High | 1.32e6 | 5.00e6 | 4.89e5 | 1.82e5 | 6.82e5 | 2.67e4 |

**Table 5:** Densities of women giving birth to unhealthy babies for different vertical transmission probabilities over 1,000. Low probability(*ψ* = 0.1), medium probability (*ψ* = 0.5), high probability (*ψ* = 0.9).

| Probability | Colombia | Brazil | Bolivia | El Salvador | Guatemala | P. Rico |
| --- | --- | --- | --- | --- | --- | --- |
| Low | 1.82e4 | 0.62e5 | 4.89e5 | 2,005 | 0.83e4 | 601 |
| Medium | 5.03e4 | 1.89e5 | 4.97e5 | 7,000 | 2.51e4 | 2,113 |
| High | 8.71e4 | 3.25e5 | 5.10e5 | 12,000 | 4.28e4 | 3,620 |

### S6 Appendix. Simulations of women giving birth to unhealthy babies for different fetal malformation probabilities. Low probability(*ψ* = 0.1**), medium probability (***ψ* = 0.5**), high probability (***ψ* = 0.9)

**Fig 8:**
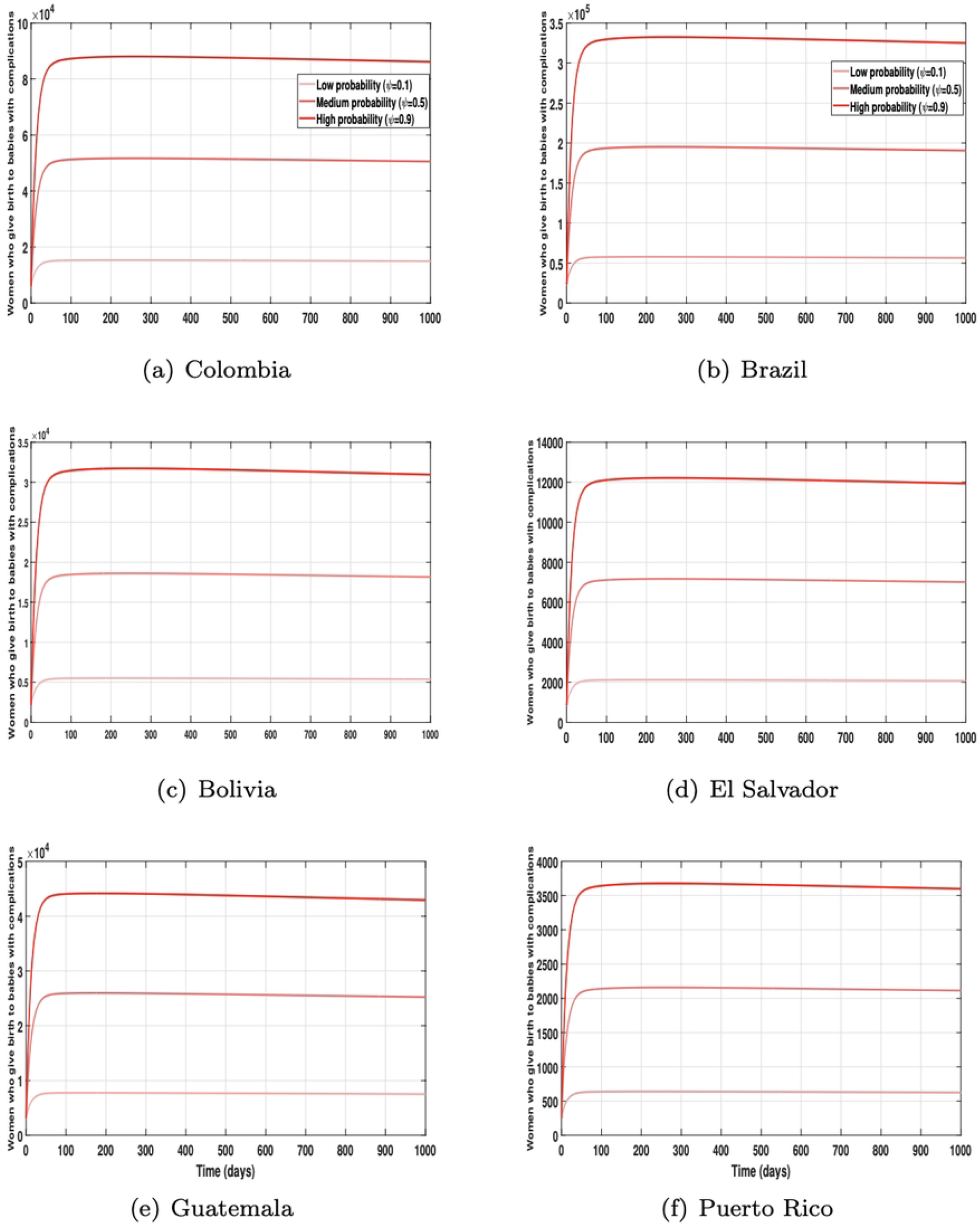
Simulations of women giving birth to unhealthy babies for different fetal malformation probabilities. Low probability(*ψ* = 0.1), medium probability (*ψ* = 0.5), high probability (*ψ* = 0.9).

### S7 Appendix. Simulations of women giving birth to healthy babies for different initial conditions for pregnant women infected with HIV *I*_*a*_(*t*)

**Fig 9:**
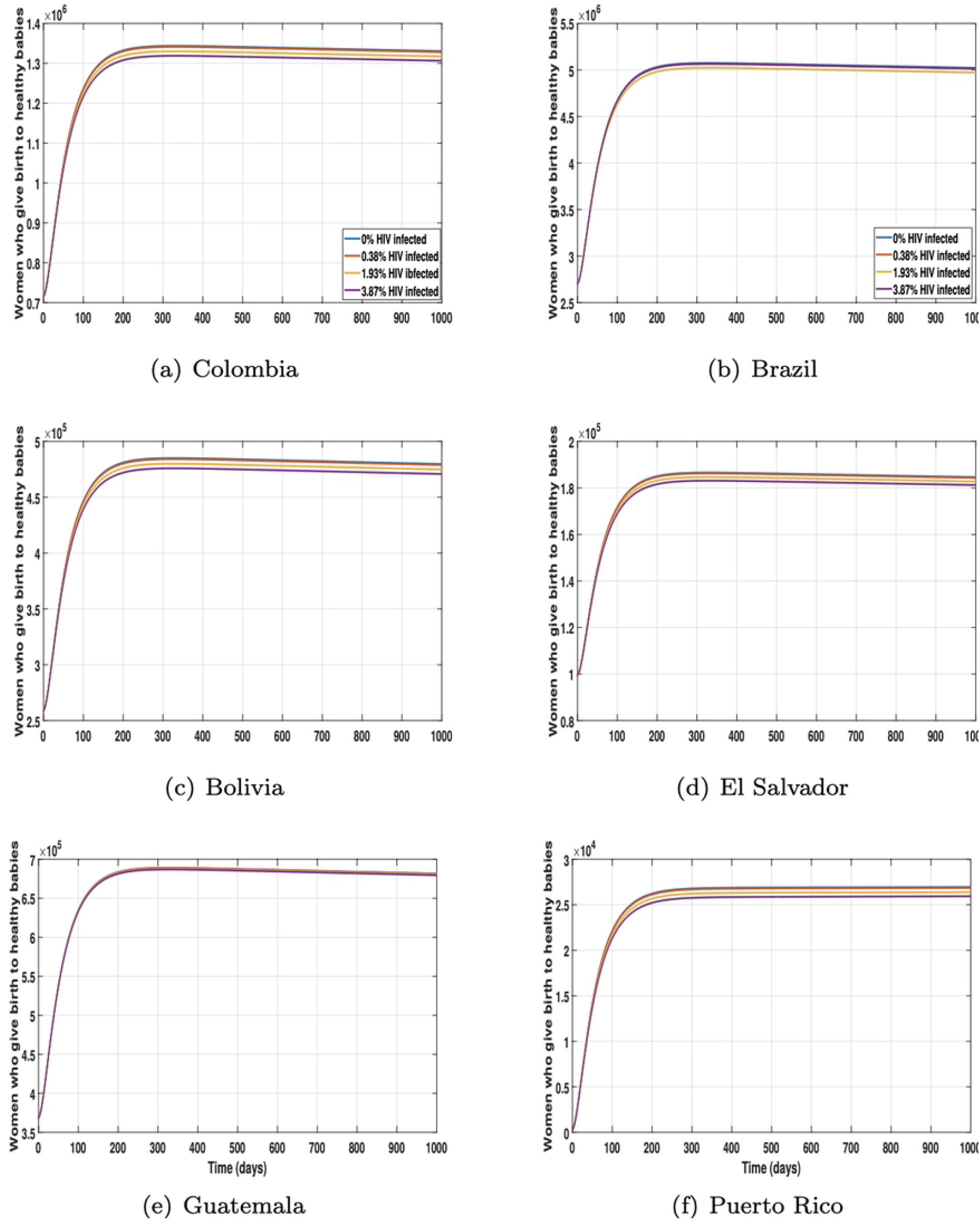
Simulations of women giving birth to healthy babies for different initial conditions for pregnant women infected with HIV *I*_*a*_(*t*).

### S8 Appendix. Simulations of women giving birth to unhealthy babies for different initial conditions for pregnant women infected with HIV *I*_*a*_(*t*)

**Fig 10:**
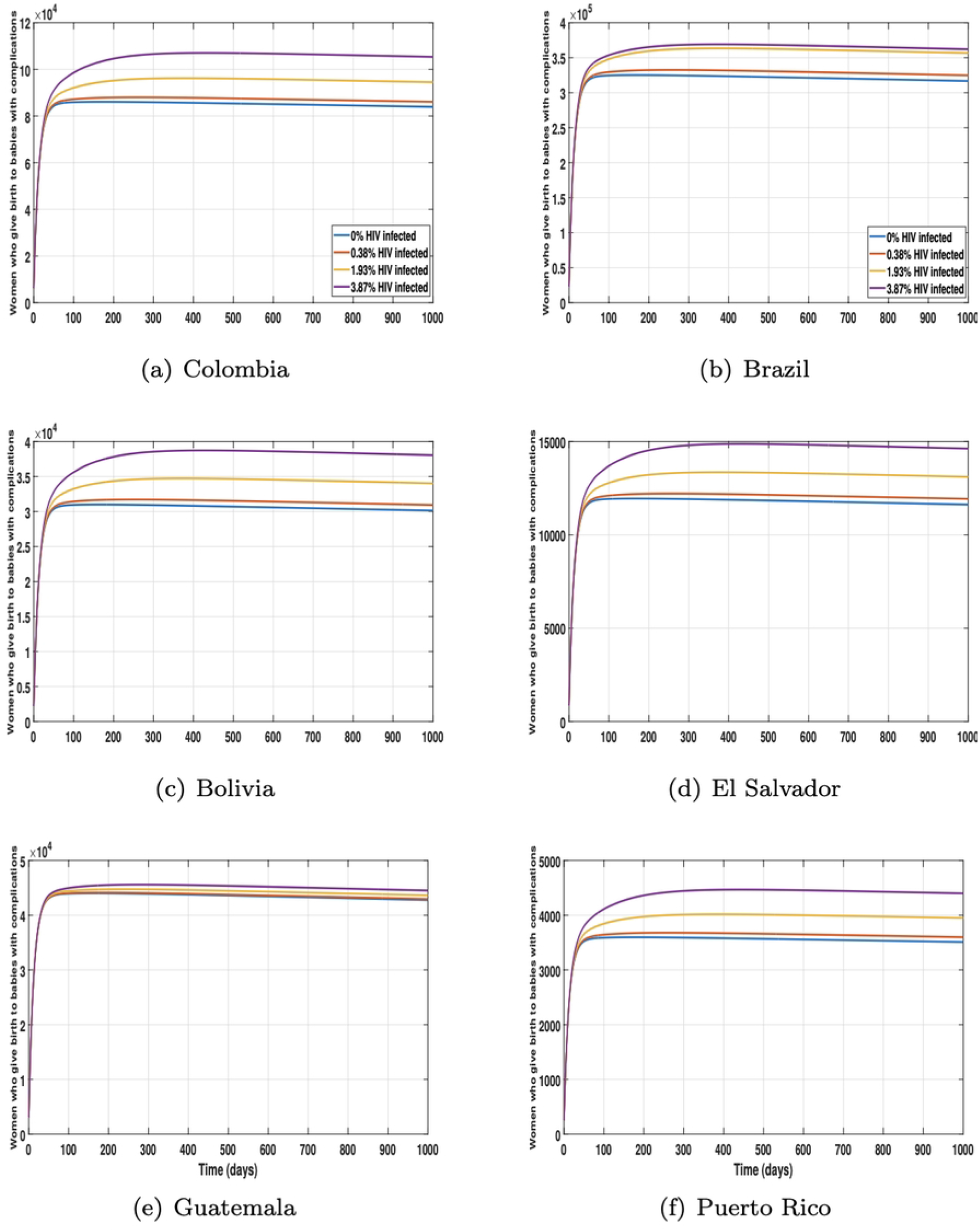
Simulations of women giving birth to unhealthy babies for different initial conditions for pregnant women infected with HIV *I*_*a*_(*t*).

**Table 6:**
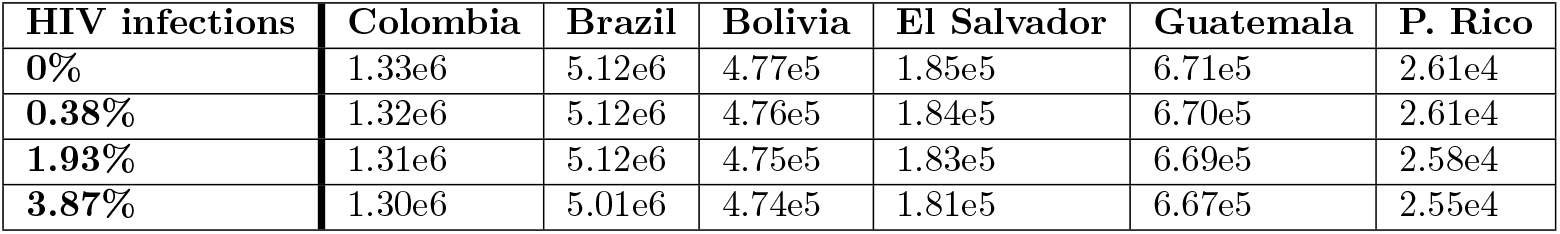
Densities of women giving birth to healthy babies for different initial conditions for pregnant women infected with HIV *I*_*a*_(*t*).

**Table 7:** Densities of women giving birth to unhealthy babies for different initial conditions for pregnant women infected with HIV *I*_*a*_(*t*).

| HIV infections | Colombia | Brazil | Bolivia | El Salvador | Guatemala | P. Rico |
| --- | --- | --- | --- | --- | --- | --- |
| 0% | 8.50e4 | 3.21e5 | 3.01e4 | 11,801 | 4.21e4 | 3,502 |
| 0.38% | 8.52e4 | 3.25e5 | 3.11e4 | 12,401 | 4.22e4 | 3,598 |
| 1.93% | 9.31e4 | 3.62e5 | 3.48e4 | 13,804 | 4.24e4 | 3,910 |
| 3.87% | 10.71e4 | 3.69e5 | 3.87e4 | 14,903 | 4.36e4 | 4,483 |

### S9 Appendix. Simulations of women giving birth to healthy babies for different initial conditions for pregnant women infected with zika *I*_*z*_(*t*)

**Fig 11:**
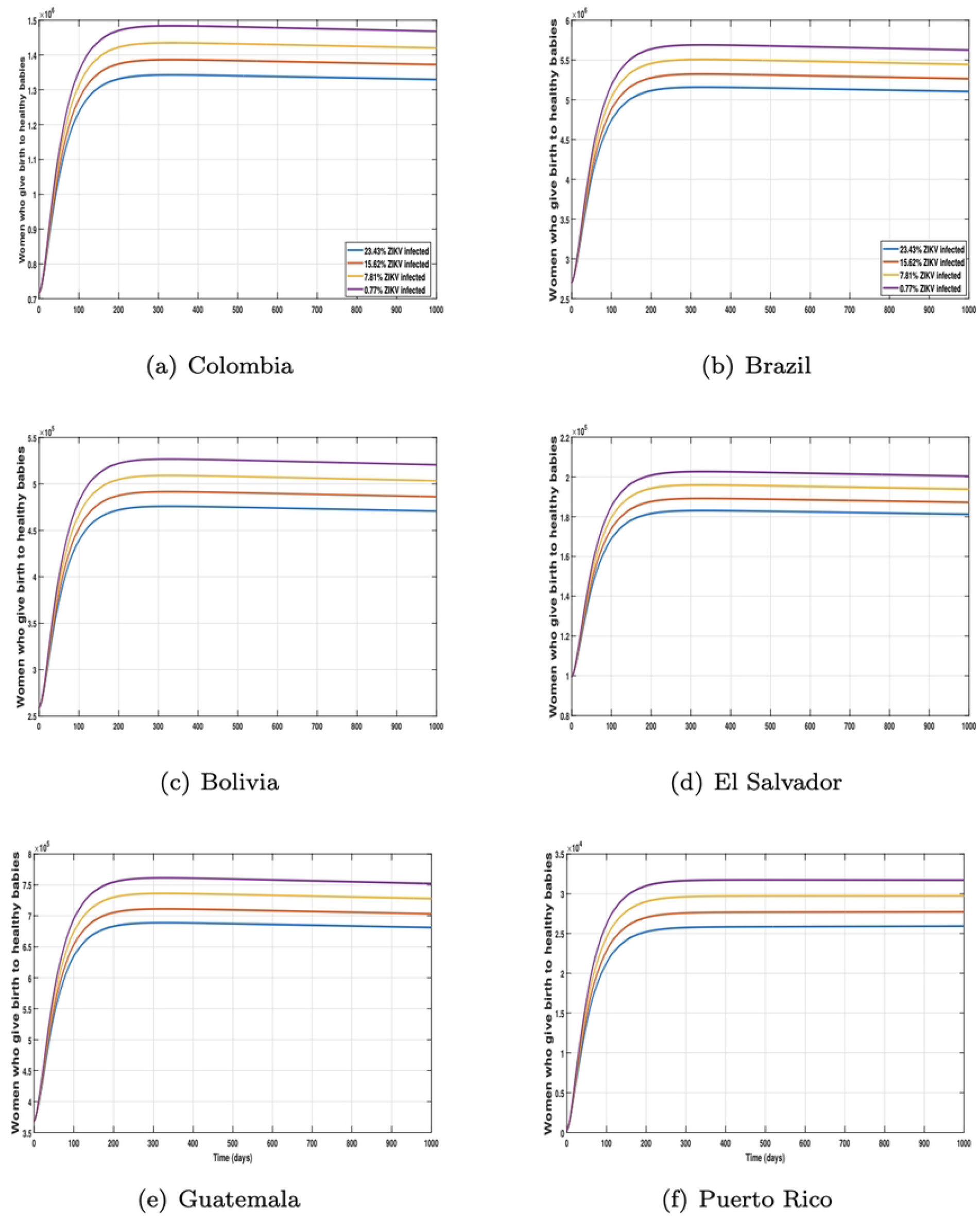
Simulations of women giving birth to healthy babies for different initial conditions for pregnant women infected with zika *I*_*z*_(*t*).

**Table 8:**
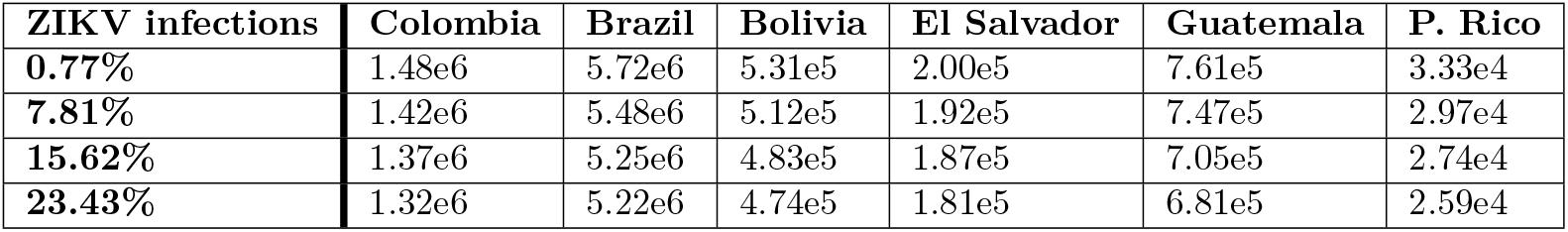
Densities of women giving birth to healthy babies for different initial conditions for pregnant women infected with zika *I*_*z*_(*t*).

**Table 9:** Densities of women giving birth to unhealthy babies for different initial condition for pregnant women infected with zika *I*_*z*_(*t*).

| ZIKV infections | Colombia | Brazil | Bolivia | El Salvador | Guatemala | P. Rico |
| --- | --- | --- | --- | --- | --- | --- |
| 0.77% | 10.50e4 | 3.71e5 | 3.89e4 | 14,320 | 4.31e4 | 4,220 |
| 7.81% | 11.50e4 | 3.92e5 | 4.00e4 | 15,603 | 4.37e4 | 4,503 |
| 15.62% | 12.00e4 | 4.27e5 | 4.27e4 | 16,504 | 4.97e4 | 4,910 |
| 23.43% | 12.85e4 | 4.48e5 | 4.49e4 | 17,505 | 5.20e4 | 5,093 |

### S10 Appendix. Simulations of women giving birth to unhealthy babies for different initial conditions for pregnant women infected with zika *I*_*z*_(*t*)

**Fig 12:**
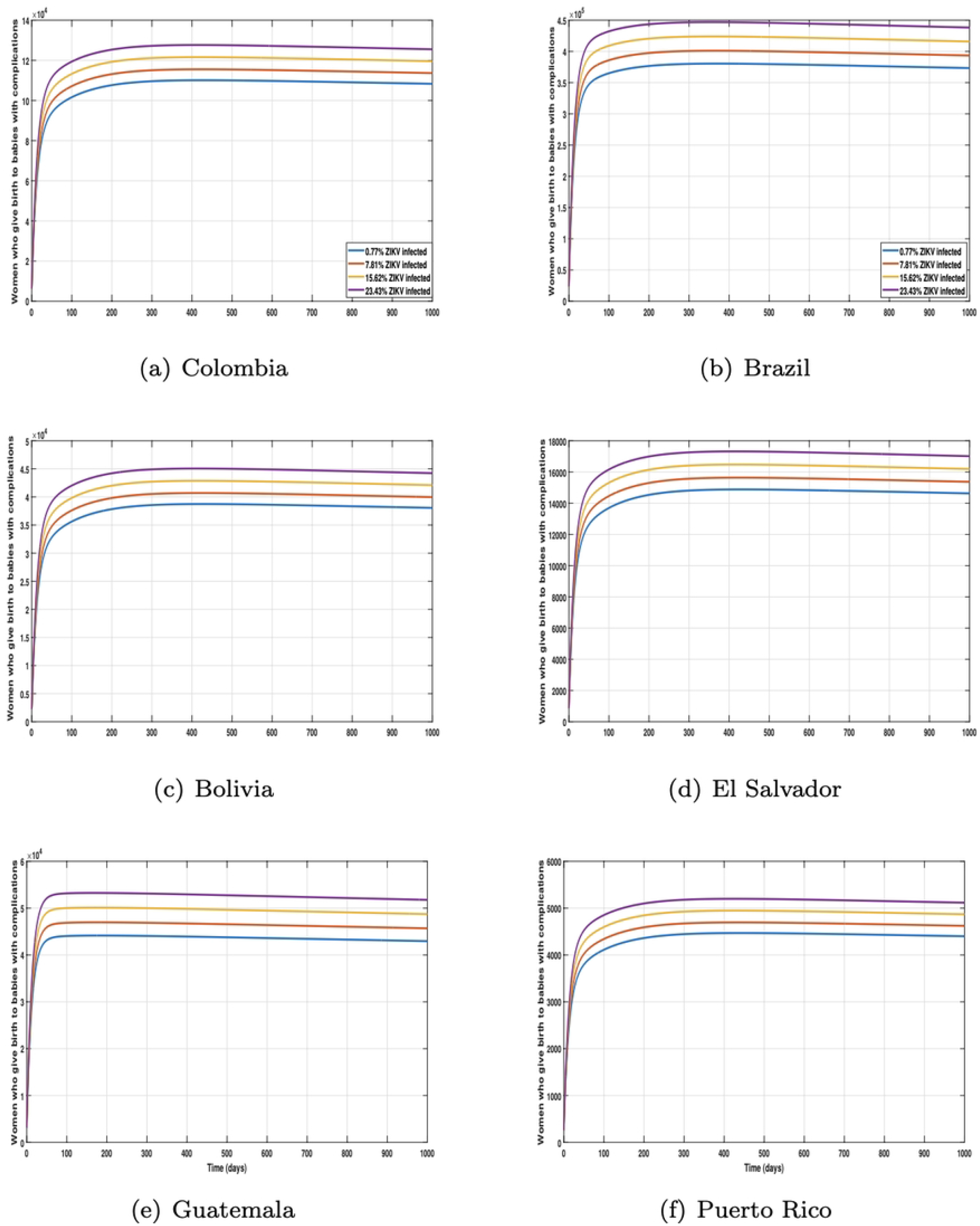
Simulations of women giving birth to unhealthy babies for different initial conditions for pregnant women infected with zika *I*_*z*_(*t*).

### S11 Appendix. Simulations of the number of birth with health issues depending on the implementation or not of control measures

Simulations depict the management of women who delivered babies with health complications. The black line illustrates the population’s behavior without control measures, while the red line reflects the population’s behavior with the activation of controls.

**Fig 13:**
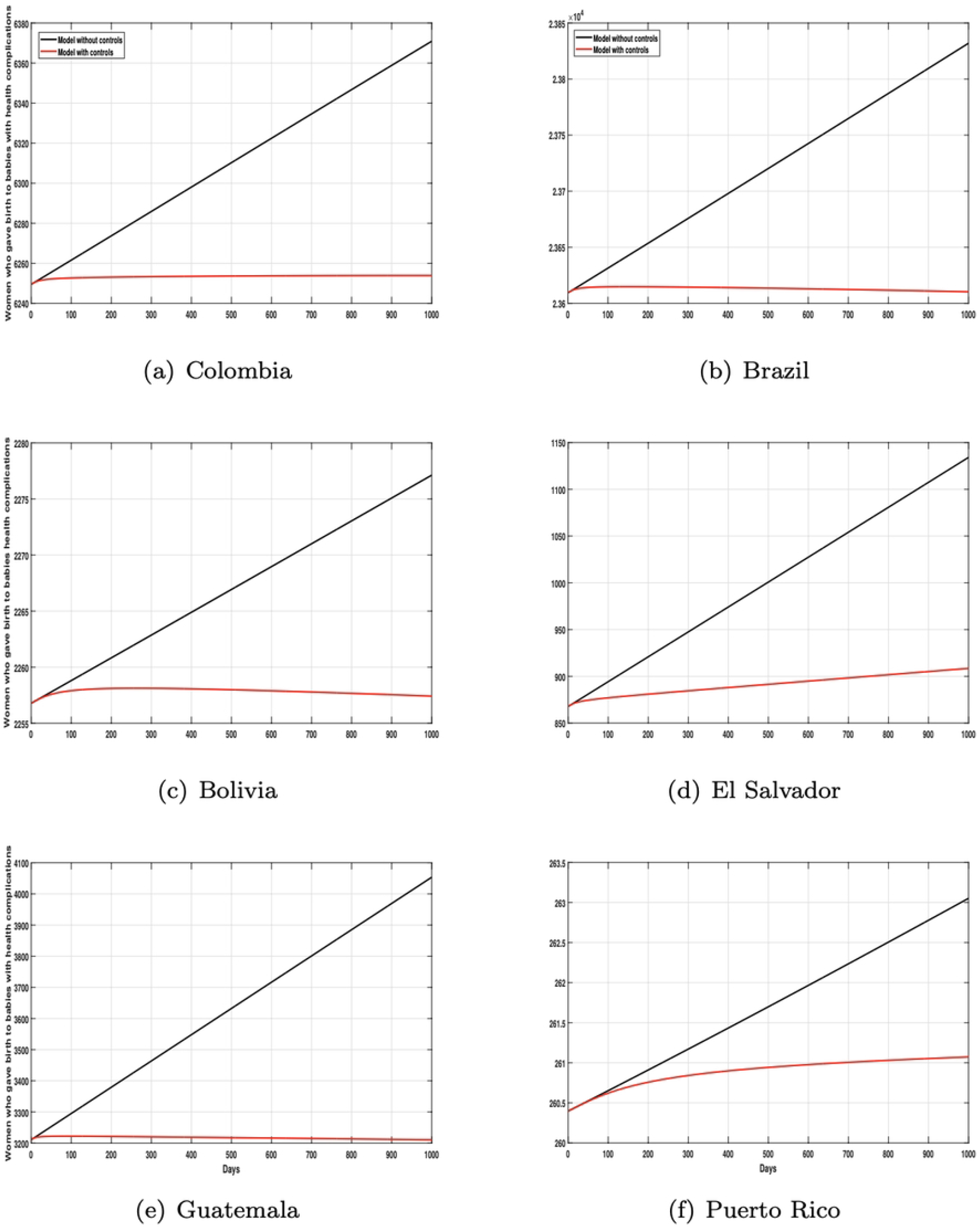
Simulations of the number of birth with health issues depending on the implementation or not of control measures.

### S12 Appendix. Projected values of the variables of interest between 2022 and 2025

The following table gives us the projected numbers for the variables of interest for the 2022-2025 period.

**Table 10:**
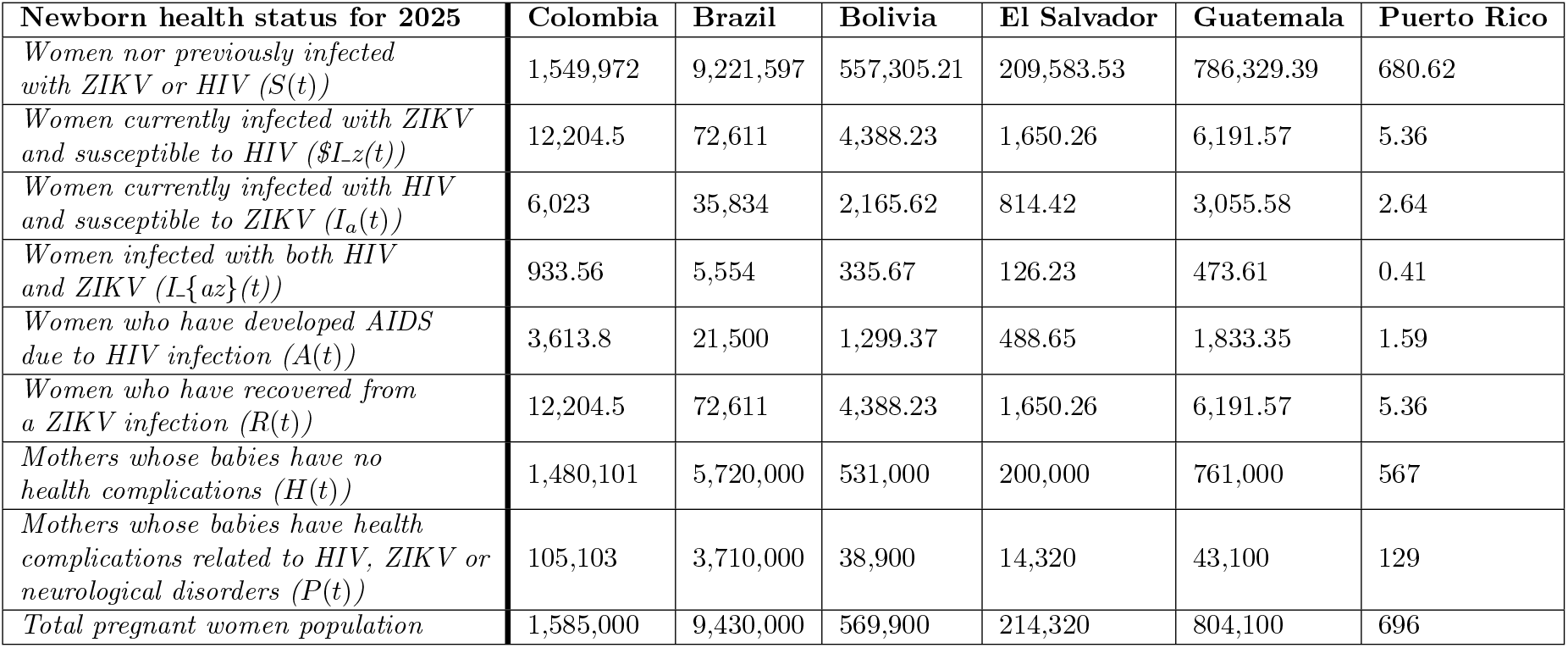
Predictions for the variables of interest for the period between 2022 and 2025.

## Acknowledgments

The authors appreciate the support provided by the One Health Modelling Network for Emerging Infections (OMNI-RÉUNIS), which is financially supported by the Natural Sciences and Engineering Research Council of Canada (NSERC) and the Public Health Agency of Canada (PHAC).

